# Optimal Allocation of COVID-19 Vaccines in the Philippines

**DOI:** 10.1101/2021.02.12.21251640

**Authors:** Christian Alvin H. Buhat, Destiny SM. Lutero, Yancee H. Olave, Kemuel M. Quindala, Mary Grace P. Recreo, Dylan Antonio SJ. Talabis, Monica C. Torres, Jerrold M. Tubay, Jomar F. Rabajante

## Abstract

Vaccine allocation is a national concern especially for countries such as the Philippines that have limited resources in acquiring COVID-19 vaccines. As such, certain groups are suggested to be prioritized for vaccination to protect the most vulnerable before vaccinating others. Our model suggests an allocation of vaccines such that COVID-19 deaths are minimized while the prioritization framework is satisfied. Results of the model show that a vaccine coverage of at least 50 to 70% of the population can be enough for a community with limited supplies, and an increase in vaccine supply is beneficial if initial coverage is less than the specified target range. Also, among the vaccines considered in the study, the one with 89.9% effectiveness and has a 183 Philippine peso (Php) price per dose projected the least number of deaths. Compared to other model variations and common allocation approaches, the model has achieved both an optimal and equitable allocation. This will be helpful for policymakers in determining a vaccine distribution for a resource-constrained community.

## 1. Introduction

The coronavirus disease 2019 (COVID-19) is a novel beta-coronavirus that was initially identified to cause respiratory problems among people from a province in China in late 2019 [1]. It is a viral infection transmitted through respiratory particles, fomites, and other biological matter [2]. Beta-coronaviruses have caused outbreaks in the recent decades, with COVID-19 being characterized as a pandemic by the World Health Organization on the first quarter of 2020 [3,4].

The Philippines is one of the countries greatly affected by this pandemic [5]. Its government placed the country under community lockdown since March 2020 as a measure to control the spread of the virus [6]. The continuous lockdown contributed to the increase in unemployment rate (10.4 percent as of December 2020) [7], decrease in remittance inflow (14-20 percent) [8], and decrease in GDP (9.5 percent decline in 2020) [9]. Bringing back jobs is crucial for a sustainable recovery of the economy [10]. All these will be possible when the COVID-19 spread is controlled.

Vaccination is considered to be an effective tool in preventing the spread of and deaths due to infectious diseases [11,12]. Vaccine development takes years to complete from clinical trials to manufacturing [13]. Vaccinating huge groups is another problem entirely. Vaccines have specific storage and shipment requirements to maintain its effectiveness [14]. Once vaccines are available for COVID-19, frontline workers should be vaccinated first followed by the elderly and people with preexisting conditions, according to various government protocols [15]. The Philippine government plans to follow the same prioritization of people for vaccination but with the addition of indigenous people and uniformed personnel in the list [16]. The plan is also to prioritize those from the cities to expedite the recovery of the local economy [17]. Distribution of vaccines will be done through local government units (LGU). Since the vaccines are limited, proper allocation of doses for each LGU is important. An optimal allocation can be found by looking at the problem as an optimization problem that minimizes the number of deaths and follows the prioritization approved by the government.

In this paper, a linear programming (LP) model is formulated to determine the optimal allocation of vaccines for every city or province in the country. The succeeding parts of the paper presents the model formulation where the parameters and assumptions of the model are discussed, followed by the results and discussion where the optimal allocation for different cases are presented together with the sensitivity analysis and parameter analysis.

## 2. Model Formulation

The LP for vaccine allocation model in this paper aims to minimize deaths while satisfying the prioritization of certain groups for vaccination as suggested by the government. The formulation of the objective function and constraints are presented. Throughout the paper, we denote the city or province where the individuals are residing as locality *i*.

### 2.1. Objective Function Formulation

The objective function value is interpreted as the projected additional COVID-19 deaths after the rollout of vaccines. It is formulated as a summation of multiplicative expressions involving the following parameters.

#### 2.1.1. Estimation of number of individuals for vaccination

To estimate the number of individuals for vaccination in each locality *i*, the locality population size *Ni*, total current locality COVID-19 cases *C*_*i*_, number of individuals for vaccination *mi*, and the effectiveness rate of the vaccine *eff* are needed. The estimate is given by

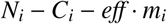

In the model, *Ni* is based on the forecasted 2020 population of the Philippines from the 2015 Philippine Statistics Authority Census [18] while *C_i_* is the total COVID-19 cases in the Philippines (as of 10 November 2020) from the DOH Data Drop [19]. For the decision variable *m_i_*, individuals that have already been previously infected are assumed to either be expired cases or recovered cases. For the recovered cases, they obtain a certain degree of immunity to COVID-19 [20]. This suggests that infected individuals can be excluded from the individuals for vaccination, which should be less than *N_i_* − *C_i_*, to further stretch the distribution of vaccines in many localities [21].

#### 2.1.2. Determination of maximum outbreak size

The maximum outbreak size [22] is computed by multiplying the number of individuals susceptible to the virus to

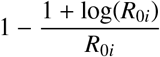

where *R*_0*i*_ is the maximum reproduction number recorded per locality *i*.

In the model, the maximum reproduction number recorded per locality *i* as of 11 November 2020 from [23] are used. Note that some localities recorded very high maximum reproduction number despite its small number of cases due to its small population [24]. Thus, we set max{*R*_0*i*_} = 4 to be the maximum reproduction number to be considered in this study [25].

#### 2.1.3. Inclusion of population density

A scaling contact coefficient described as

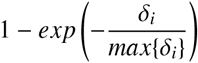

is included to incorporate the contact rate with effect of interventions, and population density *δ_i_* of each locality *i* [26]. The coefficient gives the probability of contact in the population, which is nonlinearly proportional to the population density in that locality. This coefficient characterizes the heterogeneous distribution of cases in spatial scale, which relaxes the well-mixing (homogeneity) assumption in the epidemiological model. [27, 28]. This heterogeneity in infected cases as well as deaths can be observed in Philippine setting as shown in our data dashboard (https://datastudio.google.com/s/uCv8ZX2rI8w).

### 2.2. Constraints Formulation

The following are parameters needed for the constraints of the LP model:

#### 2.2.1. Prioritization of certain groups

Due to the initial limited amount of vaccines that the country can obtain, priority groups were set by the Philippine government. These include the frontline health workers and uniformed personnel, due to their higher risk of exposure while on duty, and the senior citizens and indigent population, due to their vulnerability and by the principle of equity [29, 21]. To cater the priority groups set by the government, *G_i_* is set to be the number of individuals belonging to the priority group per locality *i*. This is set to be the minimum number of individuals for vaccination per locality.

#### 2.2.2. Incorporation of vaccination cost

A crucial factor for resource-constrained countries is the vaccination expense [30]. Some countries are considering multiple vaccines. Total expenses should be less than or equal to the allotted budget *B*. In the Philippines, the approved national budget for vaccines is 72.5 billion Philippine pesos (PHP) [31] which will cover vaccine dosage costs as well as cost of training per vaccinator (1200 PHP) and cost of other peripherals such as masks, face shield, alcohol and cotton balls (1924 PHP for 2 doses), both of which are good for 350 people [32]. The total vaccination cost is defined as

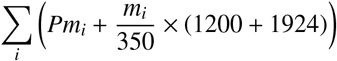

where *P* is the price per complete dose of the COVID-19 vaccine.

### 2.3. Linear Programming model

The linear programming (LP) model formulated to determine the optimal distribution of vaccines among localities in the Philippines is described below.

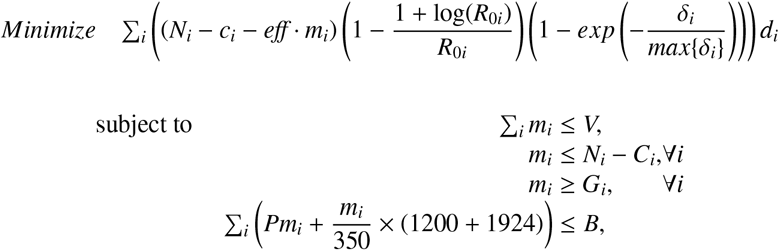

where *d*_*i*_ is the COVID-19 fatality rate per locality *i*, and *V* is the total available complete doses of vaccine for allocation across the country. The aim of this model is to find an optimal allocation that minimizes the projected number of additional COVID-19 deaths in each locality *i* such that (i) the total number of vaccines for allocation in each locality *i* will not exceed the total number of available vaccines, (ii) the individuals for vaccination per locality *i* will not include the number of individuals that have already been infected, (iii) all members of the priority groups in each locality *i* will be vaccinated, and (iv) the total cost of vaccines that will be used will not exceed the budget allocation.

## 3. Results

The LP model was used to compute for the optimal allocation under different approaches that minimizes COVID-19 deaths while satisfying the prioritization set by the government. Sensitivity analysis, effects of changes in levels of vaccine supply, vaccine effectiveness, and costs of vaccine were also studied.

### 3.1. Optimal Vaccine Allocation and Sensitivity Analysis

Budget is a huge consideration in a vaccination program so it is only fitting to study the effects of budget in vaccine allocation. The approaches where budget is considered (limited supplies and budget) and when it is not (limited supplies only) are explored in this section, with the latter approach being the basis for Sections 3.2 and 3.3. For both approaches, it is assumed that the total number of vaccines for allocation is equal to 50% of the total population, and that the effectiveness rate of the vaccine is 90%, with 2379 PHP as its cost per complete dose (similar to a market-available vaccine of 90% efficacy).

Results show that the total number of complete vaccinations to be distributed across the country is 30,361,078 when budget is considered and 54,973,950 when it is not, with corresponding deaths of 17,053 and 6,795, respectively. This translates to an average of 562 and 123 deaths per million vaccines, respectively. The more realistic scenario where budget and supplies are limited is shown in Figures 1.b, 1.d. This case has less vaccine allocation ergo more deaths, hence, supporting the need for greater budget to achieve less deaths. Furthermore, notice from Figure 1 that localities with the greater share of vaccines still resulted in large projected deaths. This can be explained by the population density and fatality risk in the locality which drives the increase in projected deaths.

**Figure 1:**
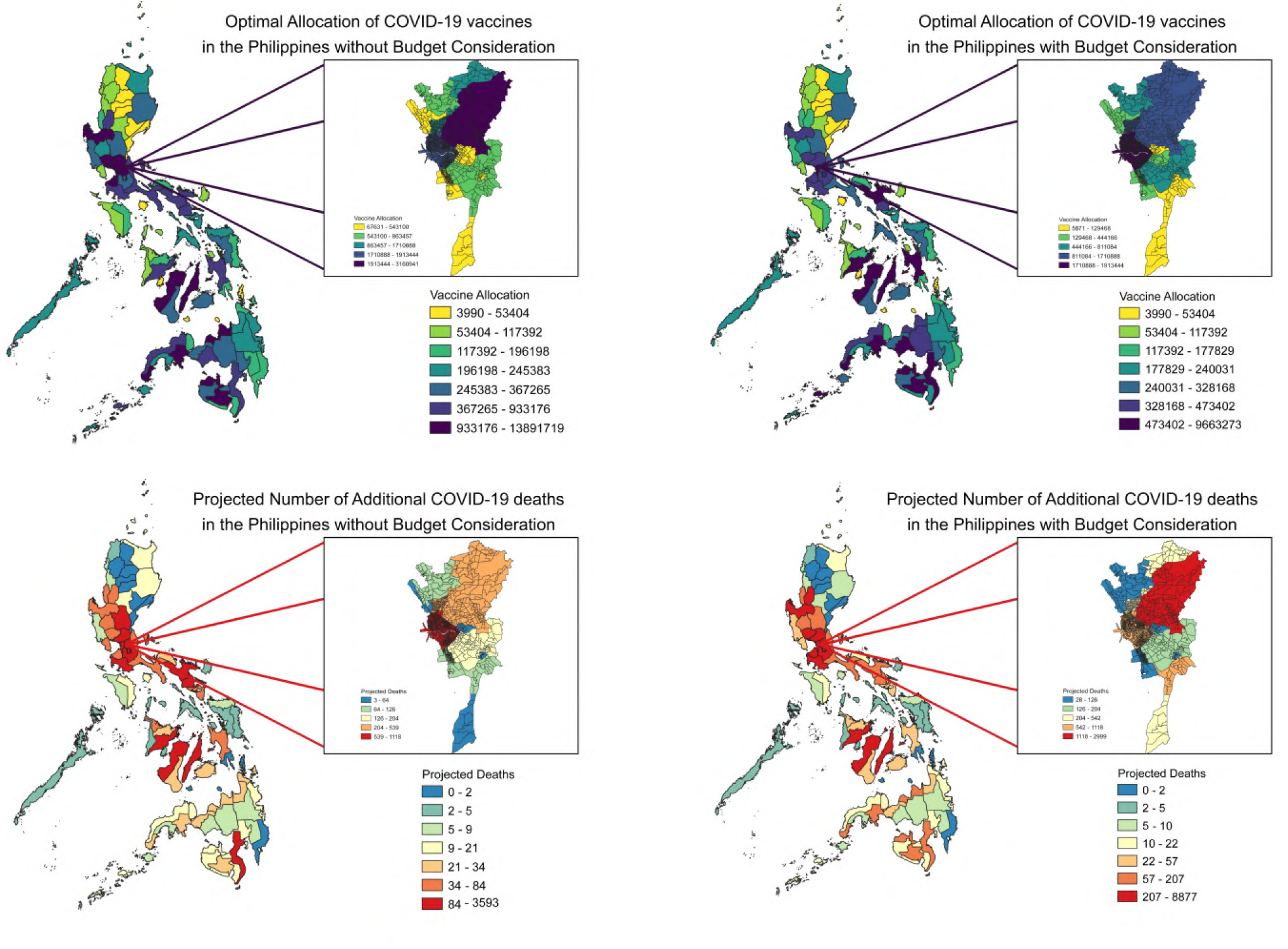
Geographical heat maps of Vaccine Allocations and its corresponding Additional Deaths (with at most 50% of the national population are vaccinated, and with 90% vaccine effectiveness rate)

Sensitivity analysis on the vaccine supply shows that, ceteris paribus, for every 1,000,000 units of increase in vaccine supply, deaths will only decrease by 112. If additional vaccines will be acquired, the best option for every unit of increase in supply should be to allocate it to Manila since for every 1000 additional vaccine allocations in Manila, projected deaths will decrease by around 5.

### 3.2. Parameter Analysis and Other Model Cases

Total vaccine supply, vaccine effectiveness, vaccine cost, and projected deaths are seen as important factors that influence vaccine allocation. Pairwise comparison of different levels of these factors are presented. Other approaches in allocating the vaccine are also studied and discussed below.

#### 3.2.1. Vaccine effectiveness and total vaccine supply levels comparison

The total vaccines to be allocated are varied from 20-100% of the total population, and the effectiveness rate of vaccines from 50-100% in the LP model without budget consideration. Note that 20% is assumed in the model to be the minimum total vaccines for allocation to accommodate all priority groups [29], and 50% to satisfy the minimum efficacy rate for COVID-19 vaccines [33].

Figure 2 shows that the greater the number of people for vaccination coupled with a high vaccine effectiveness rate, the lesser the number of additional deaths that might occur. This is an expected result. The ideal setup in a nationwide vaccination program is to maximize both factors. Unfortunately, not all countries have the resources to provide 100% coverage using the vaccine with the highest effectiveness rate. However in the heat map, notice the blue and red regions. Assume that the blue squares represent the low projected deaths and the red squares represent the high projected deaths. With low coverage of vaccination, even if there is 100% effectiveness, it will result to a red square (high number of cases). But as the coverage increases, the less minimum effectiveness necessary to achieve a blue square (low number of cases). In fact, according to the results of [34], to stop an ongoing epidemic, it is recommended that the minimum efficacy of the vaccine should be 60% when coverage is 100%, and at least 80% when coverage falls to 75%, to reduce the peak of the epidemic. Both cases fall under the blue region, among other efficacy-coverage combinations, which are feasible in the Philippines since protection and interventions will still be implemented even after vaccination [35].

**Figure 2:**
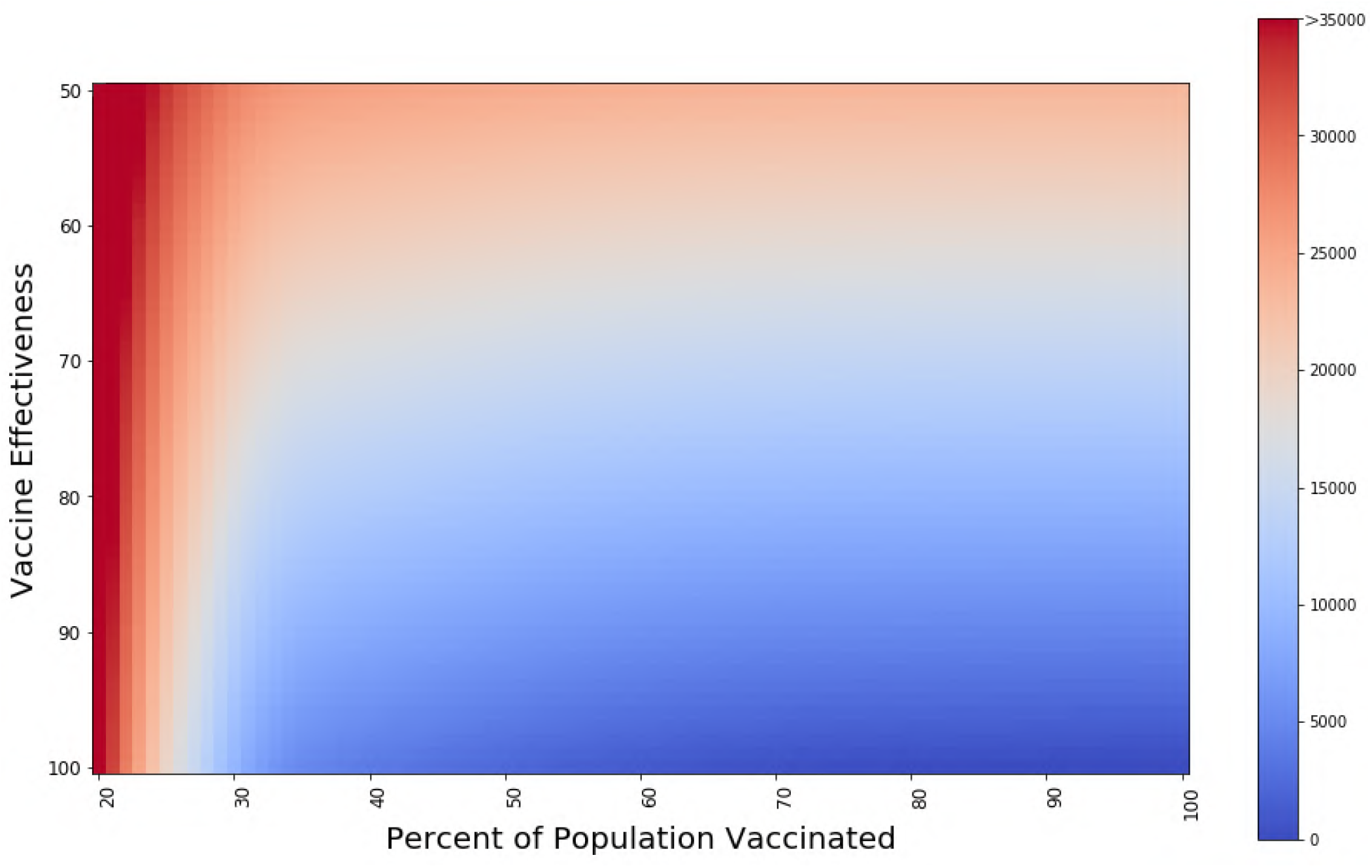
Heat map representations of the resulting objective function values (projected deaths) from the LP model without budget consideration as the percentage of population to be vaccinated is varied from 20 to 100% and the vaccine effectiveness is varied from 50 to 100%.

We further investigate the behavior of the percentage of vaccinated population and vaccine effectiveness as they increase.

From Figure 3.a, the projected deaths slowed down at around 20-40% and after 50-70%, the effect of increasing in percentage of population vaccinated is minimal. This explains the low decrease in deaths from Section 3.2, despite the huge increase in units of vaccine supply. A reason for this is since 50-70% is near the 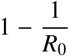 threshold for immunization coverage of the country, assuming that *R*_0_ is at a maximum [36]. This makes for a good target for countries with insufficient resources to vaccinate all of its constituents.

**Figure 3:**
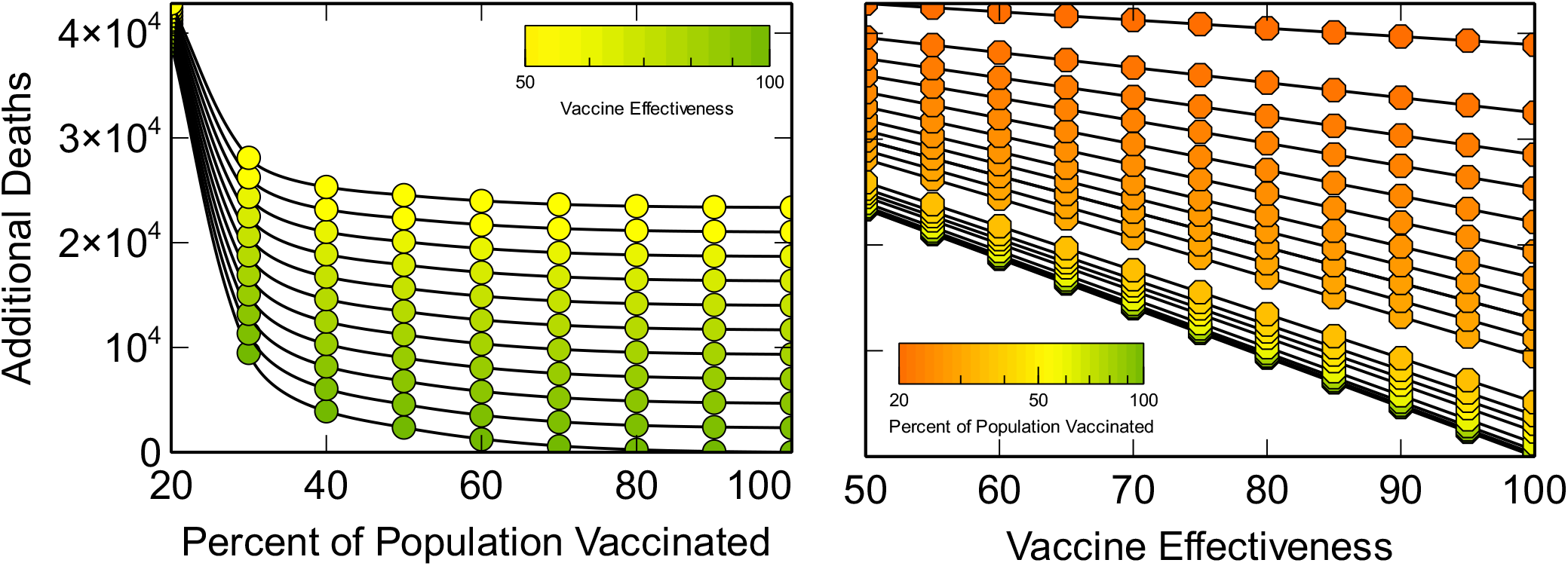
Projected additional death from the LP model as the percentage of population for vaccination and vaccine effectiveness are varied.

**Figure 4:**
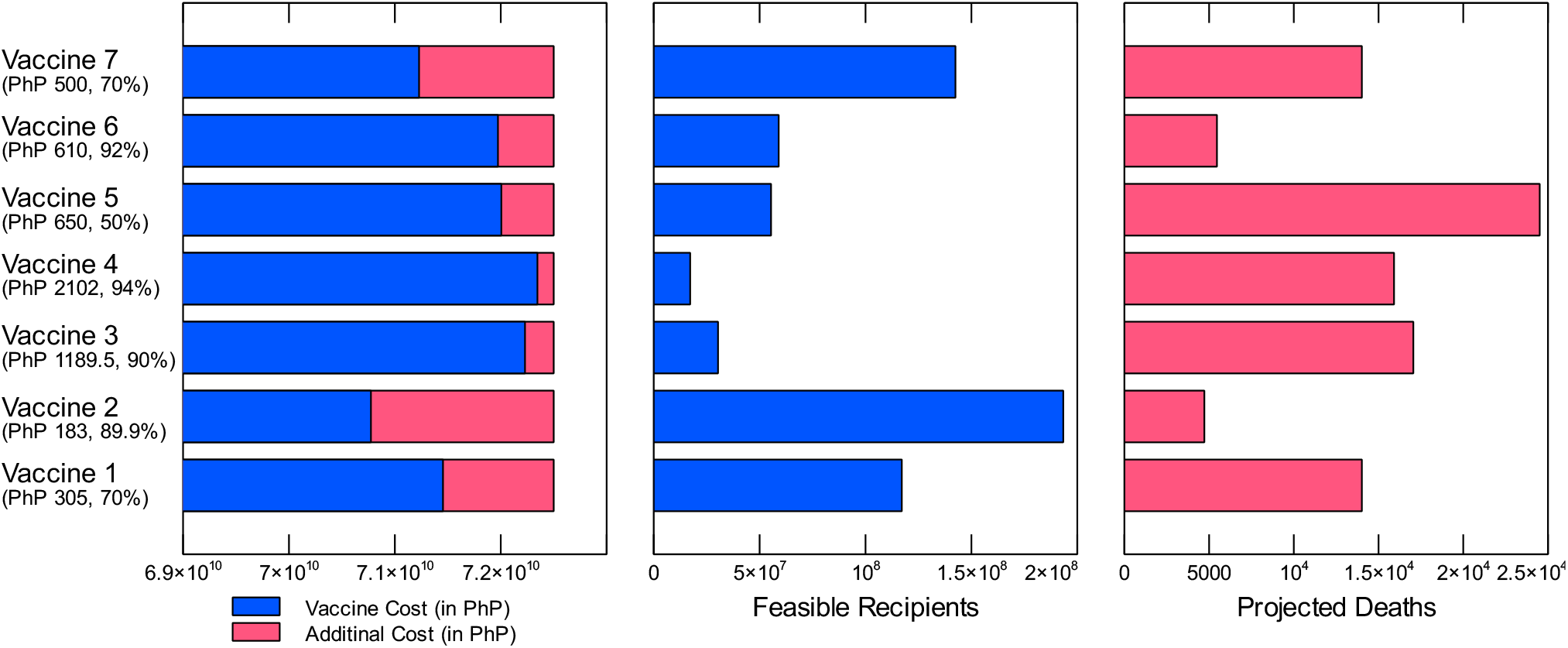
Multiple vaccines and their effectiveness, cost in PHP (1 PHP = 0.021 USD), and projected number of additional deaths based from our LP model.

Meanwhile, in Figure 3.b, the vaccine effectiveness increases have been shown to have a linear relationship with the projected deaths. That is, a vaccine with a higher effectiveness rate always generate lower deaths. Vaccine effectiveness rates should be maximized as much as possible when deciding on what vaccine should be procured. Also, notice the change in slope as the objective function value decreases, which implies that the higher the objective function value (additional deaths), the less the effect of increasing the effectiveness of the vaccine, which is derivative from the result from Figure 2 where the blue region gets larger as the number of population to be vaccinated increases, and the number of deaths decreases.

#### 3.2.2. Cost analysis in vaccine acquisition

Another key factor in determining what vaccine to use is the cost of the vaccine. Each vaccine has its own price per dose, required vaccine dose per person, and efficacy rate (as the effectiveness rate). These factors are used for comparative analysis of the vaccines.

Among the vaccines, vaccines 1, 2 and 7 can provide 100% national coverage, i.e., vaccinate all 110 million+ residents of the country. All but vaccine 4 can fulfill the prioritization of certain groups identified by the government. Vaccine 2 generated the lowest projected number of additional deaths compared to the other vaccines. This is due to its high effectiveness rate and significantly low price per dose.

Vaccines 3 and 4 can only cover 28% and 16% of the population, respectively, and have high death projections despite having high vaccine effectiveness due to its high cost per dose. Thus, a significant increase in the total budget for vaccination will drastically decrease their additional death projections (see Figure 3).

#### 3.2.3. Exploring other approaches in vaccine allocation

Multiple countries use different approaches in distributing vaccines in their community. The table below shows the other approaches for vaccine allocation that can also be adapted, including the one presented in Section 3.1.

The first approach that considers the allocated budget for vaccines presents the likely scenario in the Philippine setting, while the remaining approaches (without budget constraint), can be viewed as approaches that have equal budget allotment and are thus comparable sans budget constraint. The first two cases in Table 1 are those presented in Section 3.1. The third approach assumes that the *R*0 in every locality *i* is 4 (maximum) to further estimate what to expect if *R*0 gets bigger. As expected, there is greater projected deaths even with slight changes in the allocation per locality. If the lower bound is removed as in the fourth approach, it would result in even less deaths. However, this is unequitable since many localities will not receive vaccines.

**Table 1:** Different approaches to vaccine allocation and its projected number of deaths. Last 4 rows are without budget consideration.

| Vaccine allocation approaches | Total number of complete vaccinations | Projected number of deaths |
| --- | --- | --- |
| LP Model result<br>WITH budget consideration | 30,361,078 | 17,053 |
| LP Model result<br>WITHOUT budget consideration | 54,973,950 | 6,795 |
| LP Model result<br>WITHOUT budget consideration<br>and with $R_0 = 4$ | 54,973,950 | 8,482 |
| LP Model result<br>WITHOUT budget consideration<br>and WITHOUT lower bound | 54,973,950 | 5,900 |
| Allocation proportional<br>to population size | 54,973,949 | 25,457 |
| Allocation proportional<br>to population density | 19,952,176 | 12,588 |
| Allocation proportional<br>to number of infections | 42,391,915 | 8,433 |
| Equal allocation | 54,973,950 | 25,955 |

The next three approaches follow allocations based on proportionality, say, if a country will allocate the vaccine based on the population size of the locality, then the vaccine allocation will be proportional to the population of that locality. Note that allocations per locality cannot exceed the *N*_*i*_− *C*_*i*_ susceptible population of that province. Allocating vaccines based on population size resulted to the most number of death projections, while allocating based on the number of infections generated the least. None among the three proportion allocations had better results compared to the LP model result without budget consideration.

The last approach assumes equal allocation among localities regardless of factors considered in the previous three approaches. This, in turn, results in the most number of additional deaths, and thus, is the least ideal way to allocate the vaccines since it does not consider the difference in fatality rates, infections, dynamics on each locality.

Comparing the budget-constrained approaches, the LP model result in the second approach is the best in terms of minimizing the number of deaths and equitable considerations.

## 4. Discussion

COVID-19 vaccine allocation is an immediate concern globally. An LP model is used to study vaccine allocation that aims to minimize deaths while satisfying group prioritization for immediate vaccination. Various approaches were studied, all of which assumed using a vaccine with 90% effectiveness rate to be used for at most 50% of the population.

Sensitivity analysis shows that if only vaccine supply is limited and budget is not a constraint, Deaths will decrease by 112 for every 1 million vaccines. This figure is less than the average deaths per million vaccines for the non-budget constrained approach. As seen in Section 3.3, when vaccine coverage is between 50 to 70% which is the case for the non-budget constrained approach, decrease in deaths will not be as apparent as when coverage is less than the specified range. Additional vaccines beyond the optimal allocation therefore will be beneficial in decreasing deaths only if the vaccine coverage is less than 50%.

Various vaccines were also studied in terms of their associated costs and effectiveness, and determined that the vaccine with 89.9 % effectiveness and 183 PHP price per dose results to the lowest projected deaths. We then compared our result to various model variations and common allocation approaches, upon which our model achieved both optimal and a more equitable allocation.

Among the approaches that countries can adapt, the approach using our vaccine allocation model projects the least amount of deaths while upholding equitable considerations. Although proportional allocation according to number of infections can be done more conveniently, allocation will not yield minimal deaths. Our model thus can be used by policymakers since minimization of casualties is a humane priority. Furthermore, our model provides insight on the localities that should be prioritized when additional vaccine doses become available.

Using our model can help policymakers in distributing COVID-19 vaccines, especially if their resources are limited. The model can be applied not just to countries but also to communities, assuming the situations and health systems are similar. By having an optimal and equitable distribution of vaccines, resources are well-utilized and not wasted. Having sufficient data, the model can also be adapted to other similar infectious diseases, and can be used as basis in future distributions. Note however, that the study is limited to the use of one vaccine type only, which is not the case in some countries, and should be explored in future extensions of the study. It is also good to incorporate other factors such as new COVID-19 strains, dynamic herd immunity, group-specific infection rates and other dynamics, clinical trial results of vaccines, differential vaccine transportation costs and storage facilities, among other factors.

## Data Availability

All data are available upon request to the authors.

## 5. Acknowledgment

JFR is supported by the Abdus Salam International Centre for Theoretical Physics Associateship Scheme. This research is funded by the UP System through the UP Resilience Institute.

## Appendix A

**Table 2:**
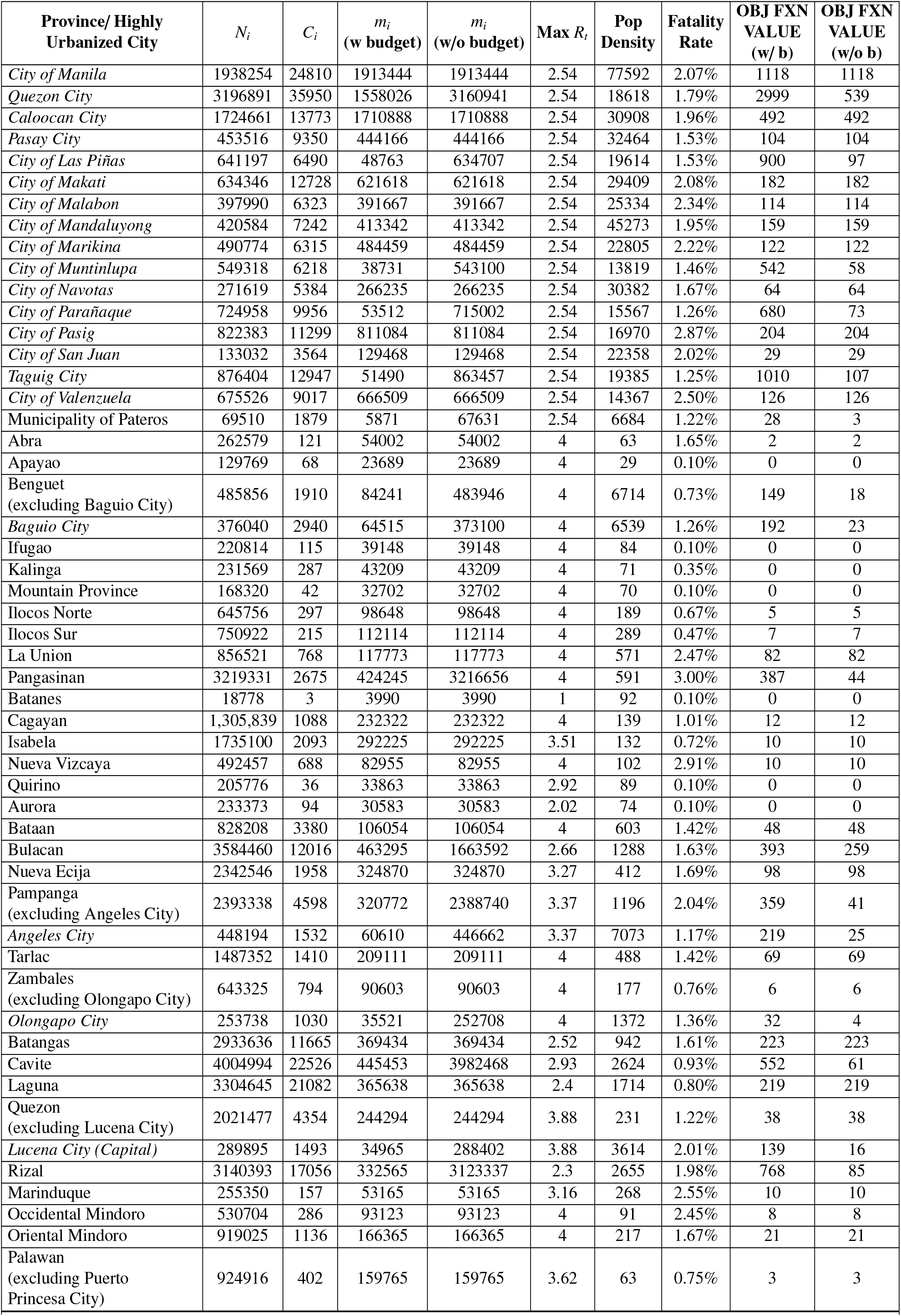
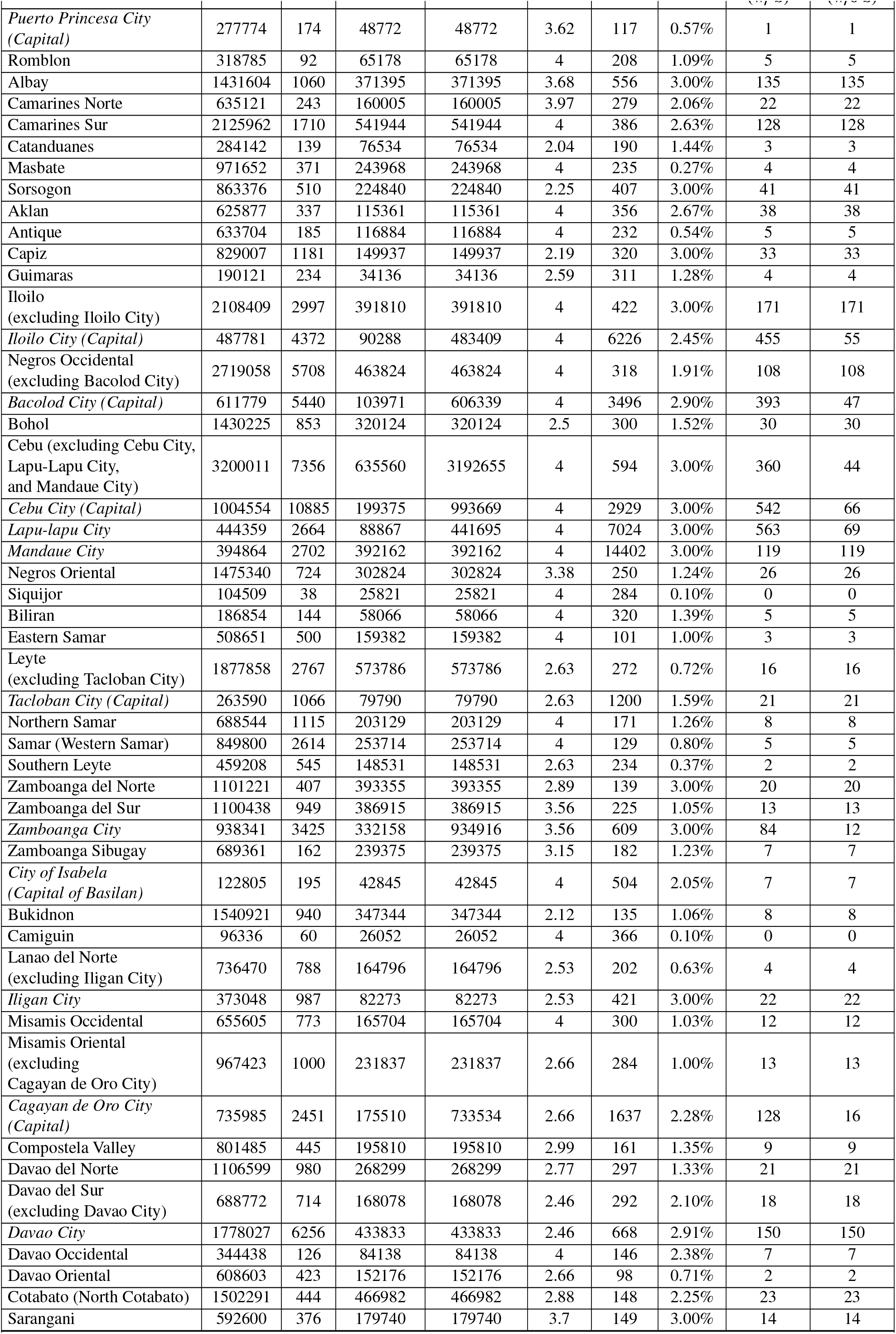
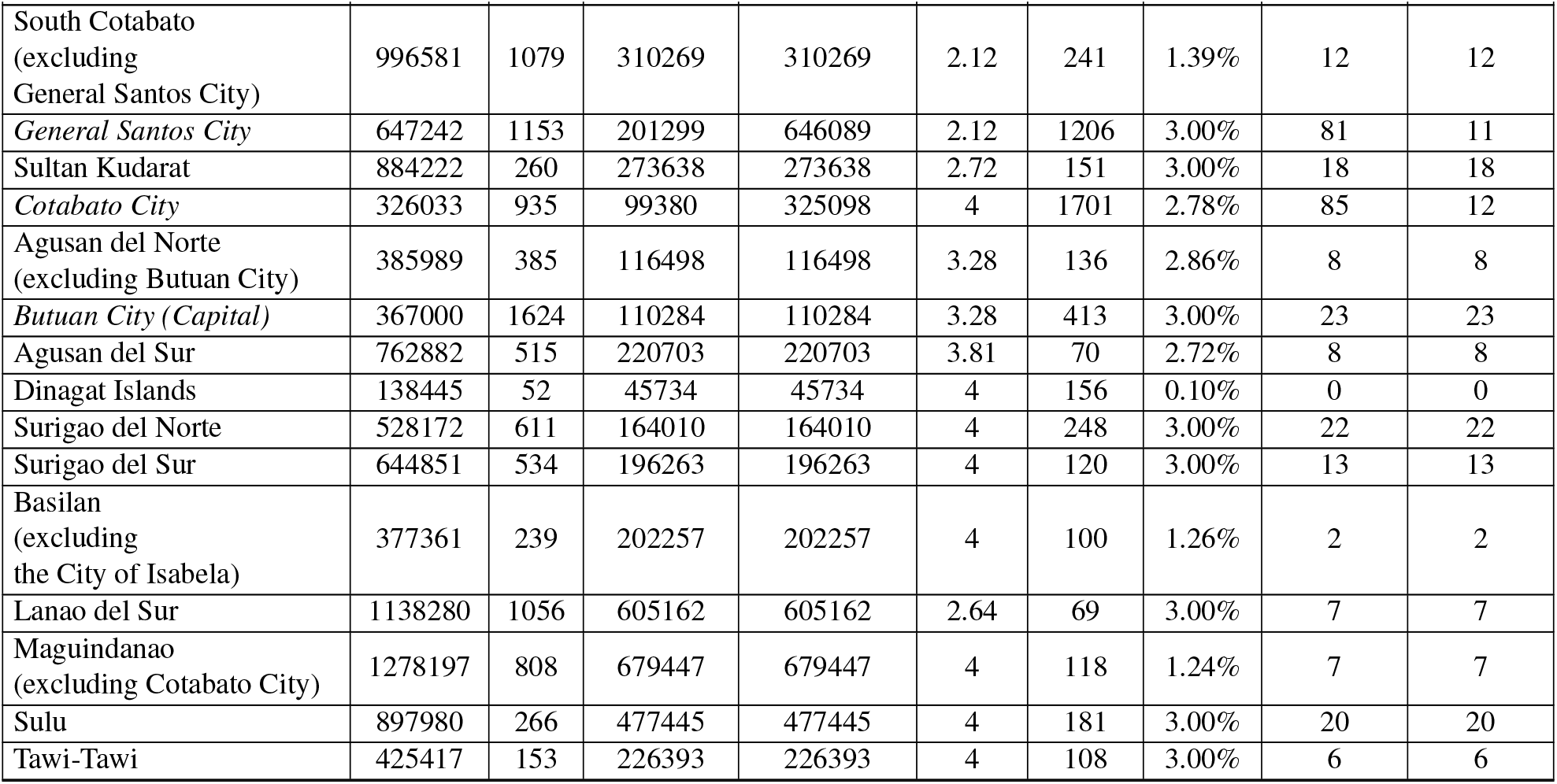
Optimal allocation of vaccines per locality with maximum 50% national vaccine coverage and 90% vaccine effectiveness

| Province/ Highly Urbanized City | $N_i$ | $C_i$ | $m_i$<br>(w budget) | $m_i$<br>(w/o budget) | Max $R_i$ | Pop Density | Fatality Rate | OBJ FXN VALUE<br>(w/ b) | OBJ FXN VALUE<br>(w/o b) |
| --- | --- | --- | --- | --- | --- | --- | --- | --- | --- |
| City of Manila | 1938254 | 24810 | 1913444 | 1913444 | 2.54 | 77592 | 2.07% | 1118 | 1118 |
| Quezon City | 3196891 | 35950 | 1558026 | 3160941 | 2.54 | 18618 | 1.79% | 2999 | 539 |
| Caloocan City | 1724661 | 13773 | 1710888 | 1710888 | 2.54 | 30908 | 1.96% | 492 | 492 |
| Pasay City | 453516 | 9350 | 444166 | 444166 | 2.54 | 32464 | 1.53% | 104 | 104 |
| City of Las Piñas | 641197 | 6490 | 48763 | 634707 | 2.54 | 19614 | 1.53% | 900 | 97 |
| City of Makati | 634346 | 12728 | 621618 | 621618 | 2.54 | 29409 | 2.08% | 182 | 182 |
| City of Malabon | 397990 | 6323 | 391667 | 391667 | 2.54 | 25334 | 2.34% | 114 | 114 |
| City of Mandaluyong | 420584 | 7242 | 413342 | 413342 | 2.54 | 45273 | 1.95% | 159 | 159 |
| City of Marikina | 490774 | 6315 | 484459 | 484459 | 2.54 | 22805 | 2.22% | 122 | 122 |
| City of Muntinlupa | 549318 | 6218 | 38731 | 543100 | 2.54 | 13819 | 1.46% | 542 | 58 |
| City of Navotas | 271619 | 5384 | 266235 | 266235 | 2.54 | 30382 | 1.67% | 64 | 64 |
| City of Parañaque | 724958 | 9956 | 53512 | 715002 | 2.54 | 15567 | 1.26% | 680 | 73 |
| City of Pasig | 822383 | 11299 | 811084 | 811084 | 2.54 | 16970 | 2.87% | 204 | 204 |
| City of San Juan | 133032 | 3564 | 129468 | 129468 | 2.54 | 22358 | 2.02% | 29 | 29 |
| Taguig City | 876404 | 12947 | 51490 | 863457 | 2.54 | 19385 | 1.25% | 1010 | 107 |
| City of Valenzuela | 675526 | 9017 | 666509 | 666509 | 2.54 | 14367 | 2.50% | 126 | 126 |
| Municipality of Pateros | 69510 | 1879 | 5871 | 67631 | 2.54 | 6684 | 1.22% | 28 | 3 |
| Abra | 262579 | 121 | 54002 | 54002 | 4 | 63 | 1.65% | 2 | 2 |
| Apayao | 129769 | 68 | 23689 | 23689 | 4 | 29 | 0.10% | 0 | 0 |
| Benguet<br>(excluding Baguio City) | 485856 | 1910 | 84241 | 483946 | 4 | 6714 | 0.73% | 149 | 18 |
| Baguio City | 376040 | 2940 | 64515 | 373100 | 4 | 6539 | 1.26% | 192 | 23 |
| Ifugao | 220814 | 115 | 39148 | 39148 | 4 | 84 | 0.10% | 0 | 0 |
| Kalinga | 231569 | 287 | 43209 | 43209 | 4 | 71 | 0.35% | 0 | 0 |
| Mountain Province | 168320 | 42 | 32702 | 32702 | 4 | 70 | 0.10% | 0 | 0 |
| Ilocos Norte | 645756 | 297 | 98648 | 98648 | 4 | 189 | 0.67% | 5 | 5 |
| Ilocos Sur | 750922 | 215 | 112114 | 112114 | 4 | 289 | 0.47% | 7 | 7 |
| La Union | 856521 | 768 | 117773 | 117773 | 4 | 571 | 2.47% | 82 | 82 |
| Pangasinan | 3219331 | 2675 | 424245 | 3216656 | 4 | 591 | 3.00% | 387 | 44 |
| Batanes | 18778 | 3 | 3990 | 3990 | 1 | 92 | 0.10% | 0 | 0 |
| Cagayan | 1,305,839 | 1088 | 232322 | 232322 | 4 | 139 | 1.01% | 12 | 12 |
| Isabela | 1735100 | 2093 | 292225 | 292225 | 3.51 | 132 | 0.72% | 10 | 10 |
| Nueva Vizcaya | 492457 | 688 | 82955 | 82955 | 4 | 102 | 2.91% | 10 | 10 |
| Quirino | 205776 | 36 | 33863 | 33863 | 2.92 | 89 | 0.10% | 0 | 0 |
| Aurora | 233373 | 94 | 30583 | 30583 | 2.02 | 74 | 0.10% | 0 | 0 |
| Bataan | 828208 | 3380 | 106054 | 106054 | 4 | 603 | 1.42% | 48 | 48 |
| Bulacan | 3584460 | 12016 | 463295 | 1663592 | 2.66 | 1288 | 1.63% | 393 | 259 |
| Nueva Ecija | 2342546 | 1958 | 324870 | 324870 | 3.27 | 412 | 1.69% | 98 | 98 |
| Pampanga<br>(excluding Angeles City) | 2393338 | 4598 | 320772 | 2388740 | 3.37 | 1196 | 2.04% | 359 | 41 |
| Angeles City | 448194 | 1532 | 60610 | 446662 | 3.37 | 7073 | 1.17% | 219 | 25 |
| Tarlac | 1487352 | 1410 | 209111 | 209111 | 4 | 488 | 1.42% | 69 | 69 |
| Zambales<br>(excluding Olongapo City) | 643325 | 794 | 90603 | 90603 | 4 | 177 | 0.76% | 6 | 6 |
| Olongapo City | 253738 | 1030 | 35521 | 252708 | 4 | 1372 | 1.36% | 32 | 4 |
| Batangas | 2933636 | 11665 | 369434 | 369434 | 2.52 | 942 | 1.61% | 223 | 223 |
| Cavite | 4004994 | 22526 | 445453 | 3982468 | 2.93 | 2624 | 0.93% | 552 | 61 |
| Laguna | 3304645 | 21082 | 365638 | 365638 | 2.4 | 1714 | 0.80% | 219 | 219 |
| Quezon<br>(excluding Lucena City) | 2021477 | 4354 | 244294 | 244294 | 3.88 | 231 | 1.22% | 38 | 38 |
| Lucena City (Capital) | 289895 | 1493 | 34965 | 288402 | 3.88 | 3614 | 2.01% | 139 | 16 |
| Rizal | 3140393 | 17056 | 332565 | 3123337 | 2.3 | 2655 | 1.98% | 768 | 85 |
| Marinduque | 255350 | 157 | 53165 | 53165 | 3.16 | 268 | 2.55% | 10 | 10 |
| Occidental Mindoro | 530704 | 286 | 93123 | 93123 | 4 | 91 | 2.45% | 8 | 8 |
| Oriental Mindoro | 919025 | 1136 | 166365 | 166365 | 4 | 217 | 1.67% | 21 | 21 |
| Palawan<br>(excluding Puerto Princesa City) | 924916 | 402 | 159765 | 159765 | 3.62 | 63 | 0.75% | 3 | 3 |

Table 2 continued from previous page
| Province/ Highly Urbanized City | $N_i$ | $C_i$ | $m_i$<br>(w budget) | $m_i$<br>(w/o budget) | Max $R_i$ | Pop Density | Fatality Rate | OBJ FXN VALUE<br>(w/ b) | OBJ FXN VALUE<br>(w/o b) |
| --- | --- | --- | --- | --- | --- | --- | --- | --- | --- |
| *Puerto Princesa City (Capital)* | 277774 | 174 | 48772 | 48772 | 3.62 | 117 | 0.57% | 1 | 1 |
| Romblon | 318785 | 92 | 65178 | 65178 | 4 | 208 | 1.09% | 5 | 5 |
| Albay | 1431604 | 1060 | 371395 | 371395 | 3.68 | 556 | 3.00% | 135 | 135 |
| Camarines Norte | 635121 | 243 | 160005 | 160005 | 3.97 | 279 | 2.06% | 22 | 22 |
| Camarines Sur | 2125962 | 1710 | 541944 | 541944 | 4 | 386 | 2.63% | 128 | 128 |
| Catanduanes | 284142 | 139 | 76534 | 76534 | 2.04 | 190 | 1.44% | 3 | 3 |
| Masbate | 971652 | 371 | 243968 | 243968 | 4 | 235 | 0.27% | 4 | 4 |
| Sorsogon | 863376 | 510 | 224840 | 224840 | 2.25 | 407 | 3.00% | 41 | 41 |
| Aklan | 625877 | 337 | 115361 | 115361 | 4 | 356 | 2.67% | 38 | 38 |
| Antique | 633704 | 185 | 116884 | 116884 | 4 | 232 | 0.54% | 5 | 5 |
| Capiz | 829007 | 1181 | 149937 | 149937 | 2.19 | 320 | 3.00% | 33 | 33 |
| Guimaras | 190121 | 234 | 34136 | 34136 | 2.59 | 311 | 1.28% | 4 | 4 |
| Iloilo<br>(excluding Iloilo City) | 2108409 | 2997 | 391810 | 391810 | 4 | 422 | 3.00% | 171 | 171 |
| *Iloilo City (Capital)* | 487781 | 4372 | 90288 | 483409 | 4 | 6226 | 2.45% | 455 | 55 |
| Negros Occidental<br>(excluding Bacolod City) | 2719058 | 5708 | 463824 | 463824 | 4 | 318 | 1.91% | 108 | 108 |
| *Bacolod City (Capital)* | 611779 | 5440 | 103971 | 606339 | 4 | 3496 | 2.90% | 393 | 47 |
| Bohol | 1430225 | 853 | 320124 | 320124 | 2.5 | 300 | 1.52% | 30 | 30 |
| Cebu (excluding Cebu City,<br>Lapu-Lapu City,<br>and Mandaue City) | 3200011 | 7356 | 635560 | 3192655 | 4 | 594 | 3.00% | 360 | 44 |
| *Cebu City (Capital)* | 1004554 | 10885 | 199375 | 993669 | 4 | 2929 | 3.00% | 542 | 66 |
| *Lapu-lapu City* | 444359 | 2664 | 88867 | 441695 | 4 | 7024 | 3.00% | 563 | 69 |
| *Mandaue City* | 394864 | 2702 | 392162 | 392162 | 4 | 14402 | 3.00% | 119 | 119 |
| Negros Oriental | 1475340 | 724 | 302824 | 302824 | 3.38 | 250 | 1.24% | 26 | 26 |
| Siquijor | 104509 | 38 | 25821 | 25821 | 4 | 284 | 0.10% | 0 | 0 |
| Biliran | 186854 | 144 | 58066 | 58066 | 4 | 320 | 1.39% | 5 | 5 |
| Eastern Samar | 508651 | 500 | 159382 | 159382 | 4 | 101 | 1.00% | 3 | 3 |
| Leyte<br>(excluding Tacloban City) | 1877858 | 2767 | 573786 | 573786 | 2.63 | 272 | 0.72% | 16 | 16 |
| *Tacloban City (Capital)* | 263590 | 1066 | 79790 | 79790 | 2.63 | 1200 | 1.59% | 21 | 21 |
| Northern Samar | 688544 | 1115 | 203129 | 203129 | 4 | 171 | 1.26% | 8 | 8 |
| Samar (Western Samar) | 849800 | 2614 | 253714 | 253714 | 4 | 129 | 0.80% | 5 | 5 |
| Southern Leyte | 459208 | 545 | 148531 | 148531 | 2.63 | 234 | 0.37% | 2 | 2 |
| Zamboanga del Norte | 1101221 | 407 | 393355 | 393355 | 2.89 | 139 | 3.00% | 20 | 20 |
| Zamboanga del Sur | 1100438 | 949 | 386915 | 386915 | 3.56 | 225 | 1.05% | 13 | 13 |
| *Zamboanga City* | 938341 | 3425 | 332158 | 934916 | 3.56 | 609 | 3.00% | 84 | 12 |
| Zamboanga Sibugay | 689361 | 162 | 239375 | 239375 | 3.15 | 182 | 1.23% | 7 | 7 |
| *City of Isabela (Capital of Basilan)* | 122805 | 195 | 42845 | 42845 | 4 | 504 | 2.05% | 7 | 7 |
| Bukidnon | 1540921 | 940 | 347344 | 347344 | 2.12 | 135 | 1.06% | 8 | 8 |
| Camiguin | 96336 | 60 | 26052 | 26052 | 4 | 366 | 0.10% | 0 | 0 |
| Lanao del Norte<br>(excluding Iligan City) | 736470 | 788 | 164796 | 164796 | 2.53 | 202 | 0.63% | 4 | 4 |
| *Iligan City* | 373048 | 987 | 82273 | 82273 | 2.53 | 421 | 3.00% | 22 | 22 |
| Misamis Occidental | 655605 | 773 | 165704 | 165704 | 4 | 300 | 1.03% | 12 | 12 |
| Misamis Oriental<br>(excluding Cagayan de Oro City) | 967423 | 1000 | 231837 | 231837 | 2.66 | 284 | 1.00% | 13 | 13 |
| *Cagayan de Oro City (Capital)* | 735985 | 2451 | 175510 | 733534 | 2.66 | 1637 | 2.28% | 128 | 16 |
| Compostela Valley | 801485 | 445 | 195810 | 195810 | 2.99 | 161 | 1.35% | 9 | 9 |
| Davao del Norte | 1106599 | 980 | 268299 | 268299 | 2.77 | 297 | 1.33% | 21 | 21 |
| Davao del Sur<br>(excluding Davao City) | 688772 | 714 | 168078 | 168078 | 2.46 | 292 | 2.10% | 18 | 18 |
| *Davao City* | 1778027 | 6256 | 433833 | 433833 | 2.46 | 668 | 2.91% | 150 | 150 |
| Davao Occidental | 344438 | 126 | 84138 | 84138 | 4 | 146 | 2.38% | 7 | 7 |
| Davao Oriental | 608603 | 423 | 152176 | 152176 | 2.66 | 98 | 0.71% | 2 | 2 |
| Cotabato (North Cotabato) | 1502291 | 444 | 466982 | 466982 | 2.88 | 148 | 2.25% | 23 | 23 |
| Sarangani | 592600 | 376 | 179740 | 179740 | 3.7 | 149 | 3.00% | 14 | 14 |

Table 2 continued from previous page
| Province/ Highly Urbanized City | $N_i$ | $C_i$ | $m_i$<br>(w budget) | $m_i$<br>(w/o budget) | Max $R_i$ | Pop Density | Fatality Rate | OBJ FXN VALUE<br>(w/ b) | OBJ FXN VALUE<br>(w/o b) |
| --- | --- | --- | --- | --- | --- | --- | --- | --- | --- |
| South Cotabato<br>(excluding General Santos City) | 996581 | 1079 | 310269 | 310269 | 2.12 | 241 | 1.39% | 12 | 12 |
| *General Santos City* | 647242 | 1153 | 201299 | 646089 | 2.12 | 1206 | 3.00% | 81 | 11 |
| Sultan Kudarat | 884222 | 260 | 273638 | 273638 | 2.72 | 151 | 3.00% | 18 | 18 |
| *Cotabato City* | 326033 | 935 | 99380 | 325098 | 4 | 1701 | 2.78% | 85 | 12 |
| Agusan del Norte<br>(excluding Butuan City) | 385989 | 385 | 116498 | 116498 | 3.28 | 136 | 2.86% | 8 | 8 |
| *Butuan City (Capital)* | 367000 | 1624 | 110284 | 110284 | 3.28 | 413 | 3.00% | 23 | 23 |
| Agusan del Sur | 762882 | 515 | 220703 | 220703 | 3.81 | 70 | 2.72% | 8 | 8 |
| Dinagat Islands | 138445 | 52 | 45734 | 45734 | 4 | 156 | 0.10% | 0 | 0 |
| Surigao del Norte | 528172 | 611 | 164010 | 164010 | 4 | 248 | 3.00% | 22 | 22 |
| Surigao del Sur | 644851 | 534 | 196263 | 196263 | 4 | 120 | 3.00% | 13 | 13 |
| Basilan<br>(excluding the City of Isabela) | 377361 | 239 | 202257 | 202257 | 4 | 100 | 1.26% | 2 | 2 |
| Lanao del Sur | 1138280 | 1056 | 605162 | 605162 | 2.64 | 69 | 3.00% | 7 | 7 |
| Maguindanao<br>(excluding Cotabato City) | 1278197 | 808 | 679447 | 679447 | 4 | 118 | 1.24% | 7 | 7 |
| Sulu | 897980 | 266 | 477445 | 477445 | 4 | 181 | 3.00% | 20 | 20 |
| Tawi-Tawi | 425417 | 153 | 226393 | 226393 | 4 | 108 | 3.00% | 6 | 6 |

## Appendix B

**Table 3:** Sensitivity analysis for the constraints specifying the maximum number of vaccines allocated to each city/province

| **Constraint for the Lower Bound of the No. of Vaccines in [city/province]** | **Solution** | **Reduced Cost** | **Obj. Coefficient** | **Allowable Increase** | **Allowable Decrease** |
| --- | --- | --- | --- | --- | --- |
| *City of Manila* | 1913443.964 | -0.005145063 | -0.005257252 | 0.005145063 | 1E+30 |
| *Quezon City* | 3160940.638 | -0.001422541 | -0.00153473 | 0.001422541 | 1E+30 |
| *Caloocan City* | 1710887.892 | -0.002478554 | -0.002590743 | 0.002478554 | 1E+30 |
| *Pasay City* | 444165.8973 | -0.001991018 | -0.002103207 | 0.001991018 | 1E+30 |
| *City of Las Piñas* | 634707.3216 | -0.001258315 | -0.001370505 | 0.001258315 | 1E+30 |
| *City of Makati* | 621618.4901 | -0.002529648 | -0.002641838 | 0.002529648 | 1E+30 |
| *City of Malabon* | 391666.538 | -0.0025103 | -0.002622489 | 0.0025103 | 1E+30 |
| *City of Mandaluyong* | 413341.5628 | -0.003349526 | -0.003461716 | 0.003349526 | 1E+30 |
| *City of Marikina* | 484459.0985 | -0.002158634 | -0.002270824 | 0.002158634 | 1E+30 |
| *City of Muntinlupa* | 543099.5674 | -0.000848154 | -0.000960344 | 0.000848154 | 1E+30 |
| *City of Navotas* | 266235.3533 | -0.002066294 | -0.002178483 | 0.002066294 | 1E+30 |
| *City of Parañaque* | 715001.7735 | -0.000805814 | -0.000918003 | 0.000805814 | 1E+30 |
| *City of Pasig* | 811083.8686 | -0.002153593 | -0.002265783 | 0.002153593 | 1E+30 |
| *City of San Juan* | 129467.5621 | -0.001922101 | -0.00203429 | 0.001922101 | 1E+30 |
| *Taguig City* | 863457.4839 | -0.001000423 | -0.001112613 | 0.001000423 | 1E+30 |
| *City of Valenzuela* | 666508.5185 | -0.00158427 | -0.001696459 | 0.00158427 | 1E+30 |
| Municipality of Pateros | 67631.02559 | -0.000294156 | -0.000406345 | 0.000294156 | 1E+30 |
| Abra | 54002.08897 | 0.000105006 | -7.18309E-06 | 1E+30 | 0.000105006 |
| Apayao | 23688.63231 | 0.000111989 | -2.0038E-07 | 1E+30 | 0.000111989 |
| Benguet (excluding Baguio City) | 483945.9157 | -0.000215629 | -0.000327819 | 0.000215629 | 1E+30 |
| *Baguio City* | 373100.0924 | -0.000436564 | -0.000548753 | 0.000436564 | 1E+30 |
| Ifugao | 39147.96054 | 0.000111603 | -5.86173E-07 | 1E+30 | 0.000111603 |
| Kalinga | 43209.26681 | 0.000110481 | -1.70868E-06 | 1E+30 | 0.000110481 |
| Mountain Province | 32701.50396 | 0.0001117 | -4.89606E-07 | 1E+30 | 0.0001117 |
| Ilocos Norte | 98648.33711 | 0.000103356 | -8.83383E-06 | 1E+30 | 0.000103356 |
| Ilocos Sur | 112113.805 | 0.000102852 | -9.3378E-06 | 1E+30 | 0.000102852 |
| La Union | 117772.5883 | 1.42737E-05 | -9.79157E-05 | 1E+30 | 1.42737E-05 |
| Pangasinan | 3216656.139 | -1.05529E-05 | -0.000122742 | 1.05529E-05 | 1E+30 |
| Batanes | 3990.17346 | 0.000112189 | 0 | 1E+30 | 0.000112189 |
| Cagayan | 232322.0836 | 0.00010243 | -9.75953E-06 | 1E+30 | 0.00010243 |
| Isabela | 292225.4496 | 0.000106033 | -6.15671E-06 | 1E+30 | 0.000106033 |
| Nueva Vizcaya | 82955.18794 | 9.15245E-05 | -2.06648E-05 | 1E+30 | 9.15245E-05 |
| Quirino | 33862.8029 | 0.000111677 | -5.12286E-07 | 1E+30 | 0.000111677 |
| Aurora | 30583.30579 | 0.000111884 | -3.05488E-07 | 1E+30 | 0.000111884 |
| Bataan | 106053.6867 | 5.28538E-05 | -5.93356E-05 | 1E+30 | 5.28538E-05 |
| Bulacan | 1663592.185 | 0 | -0.000112189 | 1.44038E-06 | 6.93926E-06 |
| Nueva Ecija | 324870.3127 | 6.90956E-05 | -4.30938E-05 | 1E+30 | 6.90956E-05 |
| Pampanga (excluding Angeles City) | 2388739.756 | -4.16604E-05 | -0.00015385 | 4.16604E-05 | 1E+30 |
| *Angeles City* | 446661.7637 | -0.000391455 | -0.000503645 | 0.000391455 | 1E+30 |
| Tarlac | 209111.1998 | 6.41871E-05 | -4.80023E-05 | 1E+30 | 6.41871E-05 |
| Zambales (excluding Olongapo City) | 90603.17704 | 0.000102889 | -9.30078E-06 | 1E+30 | 0.000102889 |
| *Olongapo City* | 252707.725 | -1.6302E-05 | -0.000128491 | 1.6302E-05 | 1E+30 |
| Batangas | 369434.4603 | 3.45146E-05 | -7.76747E-05 | 1E+30 | 3.45146E-05 |
| Cavite | 3982467.682 | -2.64694E-05 | -0.000138659 | 2.64694E-05 | 1E+30 |
| Laguna | 365638.2307 | 4.56162E-05 | -6.65732E-05 | 1E+30 | 4.56162E-05 |
| Quezon (excluding Lucena City) | 244293.9666 | 9.29426E-05 | -1.92467E-05 | 1E+30 | 9.29426E-05 |
| *Lucena City (Capital)* | 288402.1331 | -0.000373824 | -0.000486013 | 0.000373824 | 1E+30 |
| Rizal | 3123337.054 | -0.000132594 | -0.000244784 | 0.000132594 | 1E+30 |
| Marinduque | 53164.67299 | 7.06392E-05 | -4.15501E-05 | 1E+30 | 7.06392E-05 |
| Occidental Mindoro | 93123.28137 | 9.67618E-05 | -1.54275E-05 | 1E+30 | 9.67618E-05 |
| Oriental Mindoro | 166364.9017 | 8.70069E-05 | -2.51825E-05 | 1E+30 | 8.70069E-05 |
| Palawan (excluding Puerto Princesa City) | 159765.11 | 0.000109079 | -3.11062E-06 | 1E+30 | 0.000109079 |
| *Puerto Princesa City (Capital)* | 48771.93595 | 0.000107764 | -4.42495E-06 | 1E+30 | 0.000107764 |
| Romblon | 65177.82678 | 9.6498E-05 | -1.56914E-05 | 1E+30 | 9.6498E-05 |
| Albay | 371395.2791 | 1.44038E-06 | -0.000110749 | 1E+30 | 1.44038E-06 |
| Camarines Norte | 160005.0127 | 7.25156E-05 | -3.96737E-05 | 1E+30 | 7.25156E-05 |

Table 3 continued from previous page
| Constraint for the Lower Bound of the No. of Vaccines in [city/province] | Solution | Reduced Cost | Obj. Coefficient | Allowable Increase | Allowable Decrease |
| --- | --- | --- | --- | --- | --- |
| Camarines Sur | 541943.6121 | 4.17859E-05 | -7.04035E-05 | 1E+30 | 4.17859E-05 |
| Catanduanes | 76534.14304 | 0.000100825 | -1.13642E-05 | 1E+30 | 0.000100825 |
| Masbate | 243968.2537 | 0.000107796 | -4.39373E-06 | 1E+30 | 0.000107796 |
| Sorsogon | 224840.1962 | 5.57635E-05 | -5.64259E-05 | 1E+30 | 5.57635E-05 |
| Aklan | 115361.0611 | 4.63141E-05 | -6.58753E-05 | 1E+30 | 4.63141E-05 |
| Antique | 116884.0294 | 0.00010348 | -8.70961E-06 | 1E+30 | 0.00010348 |
| Capiz | 149937.2129 | 6.91485E-05 | -4.30408E-05 | 1E+30 | 6.91485E-05 |
| Guimaras | 34135.78793 | 9.12387E-05 | -2.09507E-05 | 1E+30 | 9.12387E-05 |
| Iloilo (excluding Iloilo City) | 391810.1616 | 2.44219E-05 | -8.77675E-05 | 1E+30 | 2.44219E-05 |
| *Iloilo City (Capital)* | 483408.9428 | -0.000906024 | -0.001018213 | 0.000906024 | 1E+30 |
| Negros Occidental (excluding Bacolod City) | 463824.3354 | 7.00028E-05 | -4.21866E-05 | 1E+30 | 7.00028E-05 |
| *Bacolod City (Capital)* | 606338.597 | -0.000578225 | -0.000690415 | 0.000578225 | 1E+30 |
| Bohol | 320123.7168 | 8.88815E-05 | -2.33078E-05 | 1E+30 | 8.88815E-05 |
| Cebu (excluding Cebu City, Lapu-Lapu City, and Mandaue City) | 3192655.185 | -1.13497E-05 | -0.000123539 | 1.13497E-05 | 1E+30 |
| *Cebu City (Capital)* | 993668.7942 | -0.00048741 | -0.000599599 | 0.00048741 | 1E+30 |
| *Lapu-lapu City* | 441694.9531 | -0.001288744 | -0.001400933 | 0.001288744 | 1E+30 |
| *Mandaue City* | 392161.5467 | -0.002629875 | -0.002742064 | 0.002629875 | 1E+30 |
| Negros Oriental | 302824.1482 | 9.24819E-05 | -1.97075E-05 | 1E+30 | 9.24819E-05 |
| Siquijor | 25821.16835 | 0.000110215 | -1.97399E-06 | 1E+30 | 0.000110215 |
| Biliran | 58065.81536 | 8.13327E-05 | -3.08567E-05 | 1E+30 | 8.13327E-05 |
| Eastern Samar | 159382.0767 | 0.000105158 | -7.03088E-06 | 1E+30 | 0.000105158 |
| Leyte (excluding Tacloban City) | 573786.265 | 0.000101707 | -1.04824E-05 | 1E+30 | 0.000101707 |
| *Tacloban City (Capital)* | 79789.68948 | 1.08371E-05 | -0.000101352 | 1E+30 | 1.08371E-05 |
| Northern Samar | 203128.9881 | 9.72633E-05 | -1.4926E-05 | 1E+30 | 9.72633E-05 |
| Samar (Western Samar) | 253714.3145 | 0.000104987 | -7.20283E-06 | 1E+30 | 0.000104987 |
| Southern Leyte | 148531.0295 | 0.000107611 | -4.57803E-06 | 1E+30 | 0.000107611 |
| Zamboanga del Norte | 393355.1134 | 8.83709E-05 | -2.38185E-05 | 1E+30 | 8.83709E-05 |
| Zamboanga del Sur | 386914.8738 | 9.66696E-05 | -1.55198E-05 | 1E+30 | 9.66696E-05 |
| *Zamboanga City* | 934915.7041 | -6.93926E-06 | -0.000119129 | 6.93926E-06 | 1E+30 |
| Zamboanga Sibugay | 239374.7396 | 9.85498E-05 | -1.36396E-05 | 1E+30 | 9.85498E-05 |
| *City of Isabela (Capital of Basilan)* | 42844.71763 | 4.05162E-05 | -7.16732E-05 | 1E+30 | 4.05162E-05 |
| Bukidnon | 347344.0511 | 0.000105968 | -6.22179E-06 | 1E+30 | 0.000105968 |
| Camiguin | 26051.83314 | 0.000109647 | -2.54216E-06 | 1E+30 | 0.000109647 |
| Lanao del Norte (excluding Iligan City) | 164796.1321 | 0.000105587 | -6.60203E-06 | 1E+30 | 0.000105587 |
| *Iligan City* | 82273.08047 | 4.70796E-05 | -6.51098E-05 | 1E+30 | 4.70796E-05 |
| Misamis Occidental | 165704.0384 | 9.0637E-05 | -2.15523E-05 | 1E+30 | 9.0637E-05 |
| Misamis Oriental (excluding Cagayan de Oro City) | 231837.3312 | 9.6936E-05 | -1.52533E-05 | 1E+30 | 9.6936E-05 |
| *Cagayan de Oro City (Capital)* | 733534.3039 | 0 | -0.000199387 | 8.71974E-05 | 1E+30 |
| Compostela Valley | 195810.4178 | 9.94167E-05 | -1.27726E-05 | 1E+30 | 9.94167E-05 |
| Davao del Norte | 268298.6512 | 9.03344E-05 | -2.1855E-05 | 1E+30 | 9.03344E-05 |
| Davao del Sur (excluding Davao City) | 168078.4792 | 8.12909E-05 | -3.08984E-05 | 1E+30 | 8.12909E-05 |
| *Davao City* | 433833.4173 | 1.46116E-05 | -9.75778E-05 | 1E+30 | 1.46116E-05 |
| Davao Occidental | 84137.93553 | 8.80039E-05 | -2.41854E-05 | 1E+30 | 8.80039E-05 |
| Davao Oriental | 152176.4982 | 0.000108433 | -3.75677E-06 | 1E+30 | 0.000108433 |
| Cotabato (North Cotabato) | 466982.1978 | 9.31252E-05 | -1.90642E-05 | 1E+30 | 9.31252E-05 |
| Sarangani | 179739.8157 | 8.22584E-05 | -2.99309E-05 | 1E+30 | 8.22584E-05 |
| South Cotabato (excluding General Santos City) | 310269.1562 | 9.76484E-05 | -1.45409E-05 | 1E+30 | 9.76484E-05 |
| *General Santos City* | 646089.4291 | 0 | -0.000155906 | 4.37171E-05 | 1E+30 |
| Sultan Kudarat | 273637.512 | 8.73162E-05 | -2.48732E-05 | 1E+30 | 8.73162E-05 |
| *Cotabato City* | 325097.9424 | 0 | -0.000325391 | 0.000213202 | 1E+30 |

Table 3 continued from previous page
| Constraint for the Lower Bound of the No. of Vaccines in [city/province] | Solution | Reduced Cost | Obj. Coefficient | Allowable Increase | Allowable Decrease |
| --- | --- | --- | --- | --- | --- |
| Agusan del Norte (excluding Butuan City) | 116497.9007 | 8.80158E-05 | -2.41736E-05 | 1E+30 | 8.80158E-05 |
| *Butuan City (Capital)* | 110283.9896 | 3.51455E-05 | -7.70438E-05 | 1E+30 | 3.51455E-05 |
| Agusan del Sur | 220702.6856 | 9.92561E-05 | -1.29332E-05 | 1E+30 | 9.92561E-05 |
| Dinagat Islands | 45733.67962 | 0.000111109 | -1.08049E-06 | 1E+30 | 0.000111109 |
| Surigao del Norte | 164009.7505 | 6.04487E-05 | -5.17406E-05 | 1E+30 | 6.04487E-05 |
| Surigao del Sur | 196263.0044 | 8.71624E-05 | -2.5027E-05 | 1E+30 | 8.71624E-05 |
| Basilan (excluding the City of Isabela) | 202256.9564 | 0.000103436 | -8.75383E-06 | 1E+30 | 0.000103436 |
| Lanao del Sur | 605161.8551 | 0.000101043 | -1.11464E-05 | 1E+30 | 0.000101043 |
| Maguindanao (excluding Cotabato City) | 679446.9039 | 0.000102062 | -1.01271E-05 | 1E+30 | 0.000102062 |
| Sulu | 477445.1917 | 7.43983E-05 | -3.7791E-05 | 1E+30 | 7.43983E-05 |
| Tawi-Tawi | 226393.4462 | 8.97304E-05 | -2.24589E-05 | 1E+30 | 8.97304E-05 |

**Table 4:** Sensitivity analysis for the constraints specifying the maximum number of vaccines allocated to each city/province

| **Constraint for the Lower Bound of the No. of Vaccines in [city/province]** | **Constraint Status** | **Shadow Price** | **Max Vaccine** | **Allowable Increase** | **Allowable Decrease** |
| --- | --- | --- | --- | --- | --- |
| *City of Manila* | Binding | -0.005145063 | 1913444 | 1200297.312 | 1765248.205 |
| *Quezon City* | Binding | -0.001422541 | 3160941 | 1200297.312 | 1908852.02 |
| *Caloocan City* | Binding | -0.002478554 | 1710888 | 1200297.312 | 1597172.512 |
| *Pasay City* | Binding | -0.001991018 | 444166 | 1200297.312 | 410312.6459 |
| *City of Las Piñas* | Binding | -0.001258315 | 634707 | 1200297.312 | 585944.5765 |
| *City of Makati* | Binding | -0.002529648 | 621618 | 1200297.312 | 566549.7929 |
| *City of Malabon* | Binding | -0.0025103 | 391667 | 1200297.312 | 362335.7293 |
| *City of Mandaluyong* | Binding | -0.003349526 | 413342 | 1200297.312 | 380046.1126 |
| *City of Marikina* | Binding | -0.002158634 | 484459 | 1200297.312 | 442839.2383 |
| *City of Muntinlupa* | Binding | -0.000848154 | 543100 | 1200297.312 | 504368.8103 |
| *City of Navotas* | Binding | -0.002066294 | 266235 | 1200297.312 | 247817.4461 |
| *City of Parañaque* | Binding | -0.000805814 | 715002 | 1200297.312 | 661490.0454 |
| *City of Pasig* | Binding | -0.002153593 | 811084 | 1200297.312 | 751728.9659 |
| *City of San Juan* | Binding | -0.001922101 | 129468 | 1200297.312 | 116553.518 |
| *Taguig City* | Binding | -0.001000423 | 863457 | 1200297.312 | 811967.315 |
| *City of Valenzuela* | Binding | -0.00158427 | 666509 | 1200297.312 | 623438.2509 |
| Municipality of Pateros | Binding | -0.000294156 | 67631 | 1200297.312 | 61760.26697 |
| Abra | Not Binding | 0 | 262458 | 1E+30 | 208455.8235 |
| Apayao | Not Binding | 0 | 129701 | 1E+30 | 106012.8365 |
| Benguet (excluding Baguio City) | Binding | -0.000215629 | 483946 | 1200297.312 | 399705.2373 |
| *Baguio City* | Binding | -0.000436564 | 373100 | 1200297.312 | 308585.0417 |
| Ifugao | Not Binding | 0 | 220699 | 1E+30 | 181551.1405 |
| Kalinga | Not Binding | 0 | 231282 | 1E+30 | 188073.1606 |
| Mountain Province | Not Binding | 0 | 168278 | 1E+30 | 135576.5909 |
| Ilocos Norte | Not Binding | 0 | 645459 | 1E+30 | 546810.8579 |
| Ilocos Sur | Not Binding | 0 | 750707 | 1E+30 | 638592.8794 |
| La Union | Not Binding | 0 | 855753 | 1E+30 | 737979.9343 |
| Pangasinan | Binding | -1.05529E-05 | 3216656 | 1200297.312 | 1908852.02 |
| Batanes | Not Binding | 0 | 18775 | 1E+30 | 14784.55056 |
| Cagayan | Not Binding | 0 | 1304751 | 1E+30 | 1072428.95 |
| Isabela | Not Binding | 0 | 1733007 | 1E+30 | 1440782.012 |
| Nueva Vizcaya | Not Binding | 0 | 491769 | 1E+30 | 408814.2204 |
| Quirino | Not Binding | 0 | 205740 | 1E+30 | 171877.6577 |
| Aurora | Not Binding | 0 | 233279 | 1E+30 | 202695.2012 |
| Bataan | Not Binding | 0 | 824828 | 1E+30 | 718774.3485 |
| Bulacan | Not Binding | 0 | 3572444 | 1E+30 | 1908852.02 |
| Nueva Ecija | Not Binding | 0 | 2340588 | 1E+30 | 2015717.257 |
| Pampanga (excluding Angeles City) | Binding | -4.16604E-05 | 2388740 | 1200297.312 | 1908852.02 |
| *Angeles City* | Binding | -0.000391455 | 446662 | 1200297.312 | 386051.7355 |
| Tarlac | Not Binding | 0 | 1485942 | 1E+30 | 1276831.114 |
| Zambales (excluding Olongapo City) | Not Binding | 0 | 642531 | 1E+30 | 551927.6914 |
| *Olongapo City* | Binding | -1.6302E-05 | 252708 | 1200297.312 | 217186.6616 |
| Batangas | Not Binding | 0 | 2921971 | 1E+30 | 2552536.11 |
| Cavite | Binding | -2.64694E-05 | 3982468 | 1200297.312 | 1908852.02 |
| Laguna | Not Binding | 0 | 3283563 | 1E+30 | 2917925.101 |
| Quezon (excluding Lucena City) | Not Binding | 0 | 2017123 | 1E+30 | 1772828.574 |
| *Lucena City (Capital)* | Binding | -0.000373824 | 288402 | 1200297.312 | 253437.3683 |
| Rizal | Binding | -0.000132594 | 3123337 | 1200297.312 | 1908852.02 |
| Marinduque | Not Binding | 0 | 255193 | 1E+30 | 202028.5888 |
| Occidental Mindoro | Not Binding | 0 | 530418 | 1E+30 | 437294.9732 |
| Oriental Mindoro | Not Binding | 0 | 917889 | 1E+30 | 751524.2031 |
| Palawan (excluding Puerto Princesa City) | Not Binding | 0 | 924514 | 1E+30 | 764748.4904 |
| *Puerto Princesa City (Capital)* | Not Binding | 0 | 277600 | 1E+30 | 228828.4953 |
| Romblon | Not Binding | 0 | 318693 | 1E+30 | 253514.8663 |
| Albay | Not Binding | 0 | 1430544 | 1E+30 | 1059148.557 |
| Camarines Norte | Not Binding | 0 | 634878 | 1E+30 | 474872.6258 |
| Camarines Sur | Not Binding | 0 | 2124252 | 1E+30 | 1582307.908 |
| Catanduanes | Not Binding | 0 | 284003 | 1E+30 | 207468.6853 |

Table 4 continued from previous page
| Constraint for the Lower Bound of the No. of Vaccines in [city/province] | Constraint Status | Shadow Price | Max Vaccine | Allowable Increase | Allowable Decrease |
| --- | --- | --- | --- | --- | --- |
| Masbate | Not Binding | 0 | 971281 | 1E+30 | 727312.6929 |
| Sorsogon | Not Binding | 0 | 862866 | 1E+30 | 638025.5132 |
| Aklan | Not Binding | 0 | 625540 | 1E+30 | 510178.5279 |
| Antique | Not Binding | 0 | 633519 | 1E+30 | 516635.0592 |
| Capiz | Not Binding | 0 | 827826 | 1E+30 | 677889.0134 |
| Guimaras | Not Binding | 0 | 189887 | 1E+30 | 155751.6735 |
| Iloilo (excluding Iloilo City) | Not Binding | 0 | 2105412 | 1E+30 | 1713601.552 |
| *Iloilo City (Capital)* | Binding | -0.000906024 | 483409 | 1200297.312 | 393120.5706 |
| Negros Occidental (excluding Bacolod City) | Not Binding | 0 | 2713350 | 1E+30 | 2249525.873 |
| *Bacolod City (Capital)* | Binding | -0.000578225 | 606339 | 1200297.312 | 502367.7555 |
| Bohol | Not Binding | 0 | 1429372 | 1E+30 | 1109248.678 |
| Cebu (excluding Cebu City, Lapu-Lapu City, and Mandaue City) | Binding | -1.13497E-05 | 3192655 | 1200297.312 | 1908852.02 |
| *Cebu City (Capital)* | Binding | -0.00048741 | 993669 | 1200297.312 | 794294.2143 |
| *Lapu-lapu City* | Binding | -0.001288744 | 441695 | 1200297.312 | 352828.4071 |
| *Mandaue City* | Binding | -0.002629875 | 392162 | 1200297.312 | 313859.4929 |
| Negros Oriental | Not Binding | 0 | 1474616 | 1E+30 | 1171792.346 |
| Siquijor | Not Binding | 0 | 104471 | 1E+30 | 78649.76487 |
| Biliran | Not Binding | 0 | 186710 | 1E+30 | 128644.1087 |
| Eastern Samar | Not Binding | 0 | 508151 | 1E+30 | 348769.2948 |
| Leyte (excluding Tacloban City) | Not Binding | 0 | 1875091 | 1E+30 | 1301305.153 |
| *Tacloban City (Capital)* | Not Binding | 0 | 262524 | 1E+30 | 182734.7332 |
| Northern Samar | Not Binding | 0 | 687429 | 1E+30 | 484300.5055 |
| Samar (Western Samar) | Not Binding | 0 | 847186 | 1E+30 | 593472.0346 |
| Southern Leyte | Not Binding | 0 | 458663 | 1E+30 | 310132.1988 |
| Zamboanga del Norte | Not Binding | 0 | 1100814 | 1E+30 | 707458.9598 |
| Zamboanga del Sur | Not Binding | 0 | 1099489 | 1E+30 | 712574.3406 |
| *Zamboanga City* | Binding | -6.93926E-06 | 934916 | 1200297.312 | 602757.6155 |
| Zamboanga Sibugay | Not Binding | 0 | 689199 | 1E+30 | 449824.3662 |
| *City of Isabela (Capital of Basilan)* | Not Binding | 0 | 122610 | 1E+30 | 79765.68285 |
| Bukidnon | Not Binding | 0 | 1539981 | 1E+30 | 1192636.931 |
| Camiguin | Not Binding | 0 | 96276 | 1E+30 | 70224.44575 |
| Lanao del Norte (excluding Iligan City) | Not Binding | 0 | 735682 | 1E+30 | 570885.695 |
| *Iligan City* | Not Binding | 0 | 372061 | 1E+30 | 289787.945 |
| Misamis Occidental | Not Binding | 0 | 654832 | 1E+30 | 489127.499 |
| Misamis Oriental (excluding Cagayan de Oro City) | Not Binding | 0 | 966423 | 1E+30 | 734585.6533 |
| *Cagayan de Oro City (Capital)* | Binding | -8.71974E-05 | 733534 | 1200297.312 | 558024.2467 |
| Compostela Valley | Not Binding | 0 | 801040 | 1E+30 | 605229.8016 |
| Davao del Norte | Not Binding | 0 | 1105619 | 1E+30 | 837320.0852 |
| Davao del Sur (excluding Davao City) | Not Binding | 0 | 688058 | 1E+30 | 519979.577 |
| *Davao City* | Not Binding | 0 | 1771771 | 1E+30 | 1337937.622 |
| Davao Occidental | Not Binding | 0 | 344312 | 1E+30 | 260174.3558 |
| Davao Oriental | Not Binding | 0 | 608180 | 1E+30 | 456003.0219 |
| Cotabato (North Cotabato) | Not Binding | 0 | 1501847 | 1E+30 | 1034864.674 |
| Sarangani | Not Binding | 0 | 592224 | 1E+30 | 412484.3729 |
| South Cotabato (excluding General Santos City) | Not Binding | 0 | 995502 | 1E+30 | 685233.3259 |
| *General Santos City* | Binding | -4.37171E-05 | 646089 | 1200297.312 | 444790.7283 |
| Sultan Kudarat | Not Binding | 0 | 883962 | 1E+30 | 610324.6722 |
| *Cotabato City* | Binding | -0.000213202 | 325098 | 1200297.312 | 225718.0572 |
| Agusan del Norte (excluding Butuan City) | Not Binding | 0 | 385604 | 1E+30 | 269105.7053 |
| *Butuan City (Capital)* | Not Binding | 0 | 365376 | 1E+30 | 255091.662 |

Table 4 continued from previous page
| **Constraint for the Lower Bound**<br>**of the No. of Vaccines in [city/province]** | **Constraint**<br>**Status** | **Shadow**<br>**Price** | **Max**<br>**Vaccine** | **Allowable**<br>**Increase** | **Allowable**<br>**Decrease** |
| --- | --- | --- | --- | --- | --- |
| Agusan del Sur | Not Binding | 0 | 762367 | 1E+30 | 541664.6447 |
| Dinagat Islands | Not Binding | 0 | 138393 | 1E+30 | 92659.47662 |
| Surigao del Norte | Not Binding | 0 | 527561 | 1E+30 | 363550.9177 |
| Surigao del Sur | Not Binding | 0 | 644317 | 1E+30 | 448054.3844 |
| Basilan<br>(excluding the City of Isabela) | Not Binding | 0 | 377122 | 1E+30 | 174864.87 |
| Lanao del Sur | Not Binding | 0 | 1137224 | 1E+30 | 532062.1659 |
| Maguindanao<br>(excluding Cotabato City) | Not Binding | 0 | 1277389 | 1E+30 | 597942.353 |
| Sulu | Not Binding | 0 | 897714 | 1E+30 | 420269.2738 |
| Tawi-Tawi | Not Binding | 0 | 425264 | 1E+30 | 198870.3717 |

**Table 5:** Sensitivity analysis for the constraints specifying the minimum number of vaccines allocated to each city/province

| **Constraint for the Lower Bound of the No. of Vaccines in [city/province]** | **Constraint Status** | **Shadow Price** | **Min Vaccine** | **Allowable Increase** | **Allowable Decrease** |
| --- | --- | --- | --- | --- | --- |
| *City of Manila* | Not Binding | 0 | 148196 | 1765248.205 | 1E+30 |
| *Quezon City* | Not Binding | 0 | 223045 | 2937895.848 | 1E+30 |
| *Caloocan City* | Not Binding | 0 | 113715 | 1597172.512 | 1E+30 |
| *Pasay City* | Not Binding | 0 | 33853 | 410312.6459 | 1E+30 |
| *City of Las Piñas* | Not Binding | 0 | 48763 | 585944.5765 | 1E+30 |
| *City of Makati* | Not Binding | 0 | 55069 | 566549.7929 | 1E+30 |
| *City of Malabon* | Not Binding | 0 | 29331 | 362335.7293 | 1E+30 |
| *City of Mandaluyong* | Not Binding | 0 | 33295 | 380046.1126 | 1E+30 |
| *City of Marikina* | Not Binding | 0 | 41620 | 442839.2383 | 1E+30 |
| *City of Muntinlupa* | Not Binding | 0 | 38731 | 504368.8103 | 1E+30 |
| *City of Navotas* | Not Binding | 0 | 18418 | 247817.4461 | 1E+30 |
| *City of Parañaque* | Not Binding | 0 | 53512 | 661490.0454 | 1E+30 |
| *City of Pasig* | Not Binding | 0 | 59355 | 751728.9659 | 1E+30 |
| *City of San Juan* | Not Binding | 0 | 12914 | 116553.518 | 1E+30 |
| *Taguig City* | Not Binding | 0 | 51490 | 811967.315 | 1E+30 |
| *City of Valenzuela* | Not Binding | 0 | 43070 | 623438.2509 | 1E+30 |
| Municipality of Pateros | Not Binding | 0 | 5871 | 61760.26697 | 1E+30 |
| Abra | Binding | 0.000105006 | 54002 | 208455.8235 | 0 |
| Apayao | Binding | 0.000111989 | 23689 | 106012.8365 | 0 |
| Benguet (excluding Baguio City) | Not Binding | 0 | 84241 | 399705.2373 | 1E+30 |
| *Baguio City* | Not Binding | 0 | 64515 | 308585.0417 | 1E+30 |
| Ifugao | Binding | 0.000111603 | 39148 | 181551.1405 | 0 |
| Kalinga | Binding | 0.000110481 | 43209 | 188073.1606 | 0 |
| Mountain Province | Binding | 0.0001117 | 32702 | 135576.5909 | 0 |
| Ilocos Norte | Binding | 0.000103356 | 98648 | 546810.8579 | 0 |
| Ilocos Sur | Binding | 0.000102852 | 112114 | 638592.8794 | 0 |
| La Union | Binding | 1.42737E-05 | 117773 | 737979.9343 | 0 |
| Pangasinan | Not Binding | 0 | 424245 | 2792410.993 | 1E+30 |
| Batanes | Binding | 0.000112189 | 3990 | 14784.55056 | 0 |
| Cagayan | Binding | 0.00010243 | 232322 | 1072428.95 | 0 |
| Isabela | Binding | 0.000106033 | 292225 | 1200297.312 | 0 |
| Nueva Vizcaya | Binding | 9.15245E-05 | 82955 | 408814.2204 | 0 |
| Quirino | Binding | 0.000111677 | 33863 | 171877.6577 | 0 |
| Aurora | Binding | 0.000111884 | 30583 | 202695.2012 | 0 |
| Bataan | Binding | 5.28538E-05 | 106054 | 718774.3485 | 0 |
| Bulacan | Not Binding | 0 | 463295 | 1200297.312 | 1E+30 |
| Nueva Ecija | Binding | 6.90956E-05 | 324870 | 1200297.312 | 0 |
| Pampanga (excluding Angeles City) | Not Binding | 0 | 320772 | 2067968.123 | 1E+30 |
| *Angeles City* | Not Binding | 0 | 60610 | 386051.7355 | 1E+30 |
| Tarlac | Binding | 6.41871E-05 | 209111 | 1200297.312 | 0 |
| Zambales (excluding Olongapo City) | Binding | 0.000102889 | 90603 | 551927.6914 | 0 |
| *Olongapo City* | Not Binding | 0 | 35521 | 217186.6616 | 1E+30 |
| Batangas | Binding | 3.45146E-05 | 369434 | 1200297.312 | 5.82077E-11 |
| Cavite | Not Binding | 0 | 445453 | 3537014.392 | 1E+30 |
| Laguna | Binding | 0 | 365638 | 4.65661E-10 | 1E+30 |
| Quezon (excluding Lucena City) | Binding | 9.29426E-05 | 244294 | 1200297.312 | 0 |
| *Lucena City (Capital)* | Not Binding | 0 | 34965 | 253437.3683 | 1E+30 |
| Rizal | Not Binding | 0 | 332565 | 2790772.291 | 1E+30 |
| Marinduque | Binding | 7.06392E-05 | 53165 | 202028.5888 | 0 |
| Occidental Mindoro | Binding | 9.67618E-05 | 93123 | 437294.9732 | 0 |
| Oriental Mindoro | Binding | 8.70069E-05 | 166365 | 751524.2031 | 0 |
| Palawan (excluding Puerto Princesa City) | Binding | 0.000109079 | 159765 | 764748.4904 | 0 |
| *Puerto Princesa City (Capital)* | Binding | 0.000107764 | 48772 | 228828.4953 | 0 |
| Romblon | Binding | 9.6498E-05 | 65178 | 253514.8663 | 0 |
| Albay | Binding | 1.44038E-06 | 371395 | 1059148.557 | 0 |
| Camarines Norte | Binding | 7.25156E-05 | 160005 | 474872.6258 | 0 |
| Camarines Sur | Binding | 4.17859E-05 | 541944 | 1200297.312 | 0 |
| Catanduanes | Binding | 0.000100825 | 76534 | 207468.6853 | 0 |

Table 5 continued from previous page
| Constraint for the Lower Bound of the No. of Vaccines in [city/province] | Constraint Status | Shadow Price | Min Vaccine | Allowable Increase | Allowable Decrease |
| --- | --- | --- | --- | --- | --- |
| Masbate | Binding | 0.000107796 | 243968 | 727312.6929 | 0 |
| Sorsogon | Binding | 5.57635E-05 | 224840 | 638025.5132 | 0 |
| Aklan | Binding | 4.63141E-05 | 115361 | 510178.5279 | 0 |
| Antique | Binding | 0.00010348 | 116884 | 516635.0592 | 0 |
| Capiz | Binding | 6.91485E-05 | 149937 | 677889.0134 | 0 |
| Guimaras | Binding | 9.12387E-05 | 34136 | 155751.6735 | 0 |
| Iloilo (excluding Iloilo City) | Binding | 2.44219E-05 | 391810 | 1200297.312 | 0 |
| *Iloilo City (Capital)* | Not Binding | 0 | 90288 | 393120.5706 | 1E+30 |
| Negros Occidental (excluding Bacolod City) | Binding | 7.00028E-05 | 463824 | 1200297.312 | 0 |
| *Bacolod City (Capital)* | Not Binding | 0 | 103971 | 502367.7555 | 1E+30 |
| Bohol | Binding | 8.88815E-05 | 320124 | 1109248.678 | 0 |
| Cebu (excluding Cebu City, Lapu-Lapu City, and Mandaue City) | Not Binding | 0 | 635560 | 2557095.082 | 1E+30 |
| *Cebu City (Capital)* | Not Binding | 0 | 199375 | 794294.2143 | 1E+30 |
| *Lapu-lapu City* | Not Binding | 0 | 88867 | 352828.4071 | 1E+30 |
| *Mandaue City* | Not Binding | 0 | 78302 | 313859.4929 | 1E+30 |
| Negros Oriental | Binding | 9.24819E-05 | 302824 | 1171792.346 | 0 |
| Siquijor | Binding | 0.000110215 | 25821 | 78649.76487 | 0 |
| Biliran | Binding | 8.13327E-05 | 58066 | 128644.1087 | 0 |
| Eastern Samar | Binding | 0.000105158 | 159382 | 348769.2948 | 0 |
| Leyte (excluding Tacloban City) | Binding | 0.000101707 | 573786 | 1200297.312 | 0 |
| *Tacloban City (Capital)* | Binding | 1.08371E-05 | 79790 | 182734.7332 | 0 |
| Northern Samar | Binding | 9.72633E-05 | 203129 | 484300.5055 | 0 |
| Samar (Western Samar) | Binding | 0.000104987 | 253714 | 593472.0346 | 0 |
| Southern Leyte | Binding | 0.000107611 | 148531 | 310132.1988 | 0 |
| Zamboanga del Norte | Binding | 8.83709E-05 | 393355 | 707458.9598 | 0 |
| Zamboanga del Sur | Binding | 9.66696E-05 | 386915 | 712574.3406 | 0 |
| *Zamboanga City* | Not Binding | 0 | 332158 | 602757.6155 | 1E+30 |
| Zamboanga Sibugay | Binding | 9.85498E-05 | 239375 | 449824.3662 | 0 |
| *City of Isabela (Capital of Basilan)* | Binding | 4.05162E-05 | 42845 | 79765.68285 | 7.27596E-12 |
| Bukidnon | Binding | 0.000105968 | 347344 | 1192636.931 | 0 |
| Camiguin | Binding | 0.000109647 | 26052 | 70224.44575 | 0 |
| Lanao del Norte (excluding Iligan City) | Binding | 0.000105587 | 164796 | 570885.695 | 0 |
| *Iligan City* | Binding | 4.70796E-05 | 82273 | 289787.945 | 0 |
| Misamis Occidental | Binding | 9.0637E-05 | 165704 | 489127.499 | 0 |
| Misamis Oriental (excluding Cagayan de Oro City) | Binding | 9.6936E-05 | 231837 | 734585.6533 | 0 |
| *Cagayan de Oro City (Capital)* | Not Binding | 0 | 175510 | 558024.2467 | 1E+30 |
| Compostela Valley | Binding | 9.94167E-05 | 195810 | 605229.8016 | 0 |
| Davao del Norte | Binding | 9.03344E-05 | 268299 | 837320.0852 | 0 |
| Davao del Sur (excluding Davao City) | Binding | 8.12909E-05 | 168078 | 519979.577 | 0 |
| *Davao City* | Binding | 1.46116E-05 | 433833 | 1200297.312 | 0 |
| Davao Occidental | Binding | 8.80039E-05 | 84138 | 260174.3558 | 0 |
| Davao Oriental | Binding | 0.000108433 | 152176 | 456003.0219 | 0 |
| Cotabato (North Cotabato) | Binding | 9.31252E-05 | 466982 | 1034864.674 | 0 |
| Sarangani | Binding | 8.22584E-05 | 179740 | 412484.3729 | 0 |
| South Cotabato (excluding General Santos City) | Binding | 9.76484E-05 | 310269 | 685233.3259 | 0 |
| *General Santos City* | Not Binding | 0 | 201299 | 444790.7283 | 1E+30 |
| Sultan Kudarat | Binding | 8.73162E-05 | 273638 | 610324.6722 | 0 |
| *Cotabato City* | Not Binding | 0 | 99380 | 225718.0572 | 1E+30 |
| Agusan del Norte (excluding Butuan City) | Binding | 8.80158E-05 | 116498 | 269105.7053 | 0 |
| *Butuan City (Capital)* | Binding | 3.51455E-05 | 110284 | 255091.662 | 0 |

Table 5 continued from previous page
| Constraint for the Lower Bound of the No. of Vaccines in [city/province] | Constraint Status | Shadow Price | Min Vaccine | Allowable Increase | Allowable Decrease |
| --- | --- | --- | --- | --- | --- |
| Agusan del Sur | Binding | 9.92561E-05 | 220703 | 541664.6447 | 0 |
| Dinagat Islands | Binding | 0.000111109 | 45734 | 92659.47662 | 0 |
| Surigao del Norte | Binding | 6.04487E-05 | 164010 | 363550.9177 | 0 |
| Surigao del Sur | Binding | 8.71624E-05 | 196263 | 448054.3844 | 0 |
| Basilan<br>(excluding the City of Isabela) | Binding | 0.000103436 | 202257 | 174864.87 | 0 |
| Lanao del Sur | Binding | 0.000101043 | 605162 | 532062.1659 | 0 |
| Maguindanao<br>(excluding Cotabato City) | Binding | 0.000102062 | 679447 | 597942.353 | 0 |
| Sulu | Binding | 7.43983E-05 | 477445 | 420269.2738 | 0 |
| Tawi-Tawi | Binding | 8.97304E-05 | 226393 | 198870.3717 | 0 |

**Table 6:**
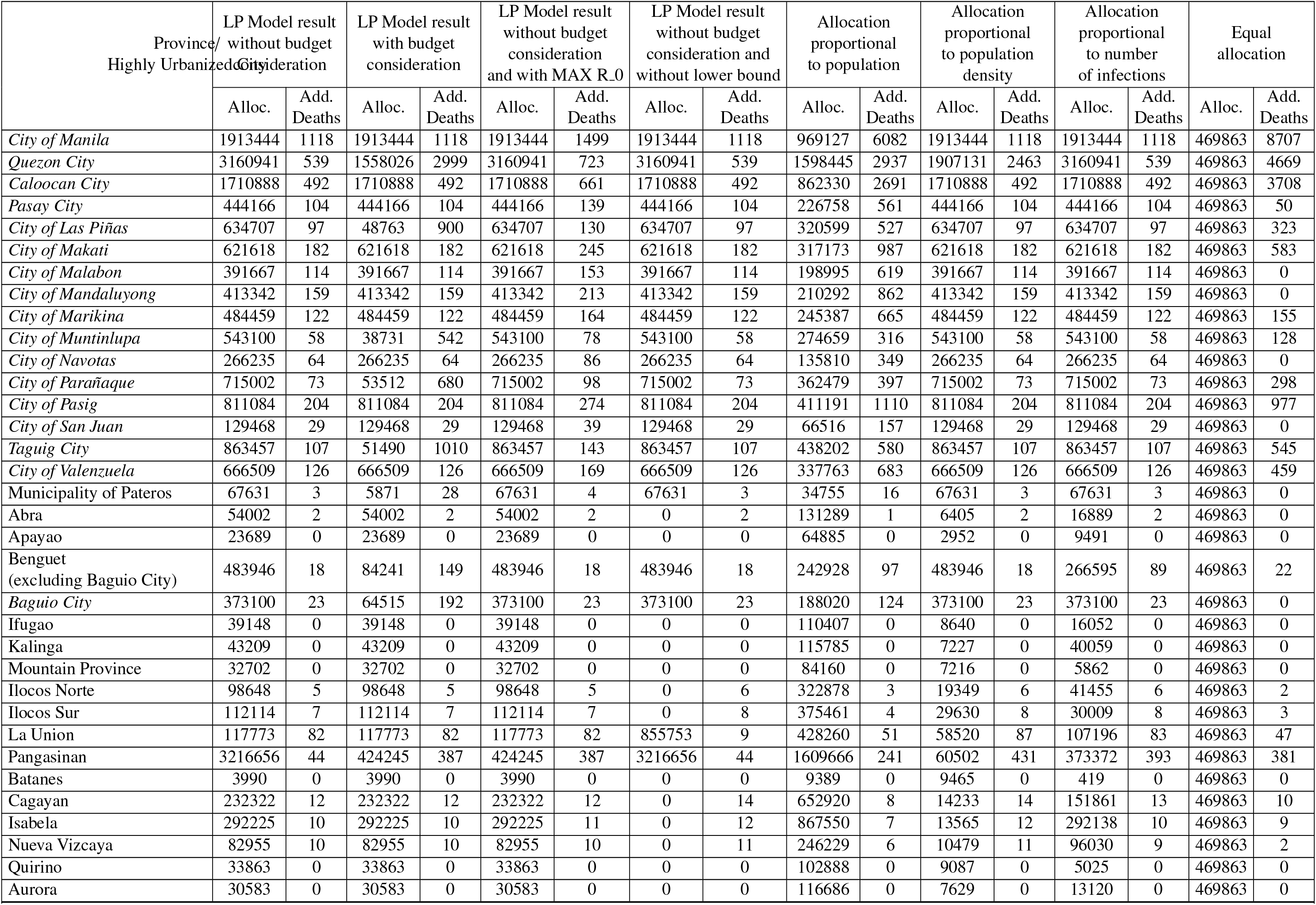
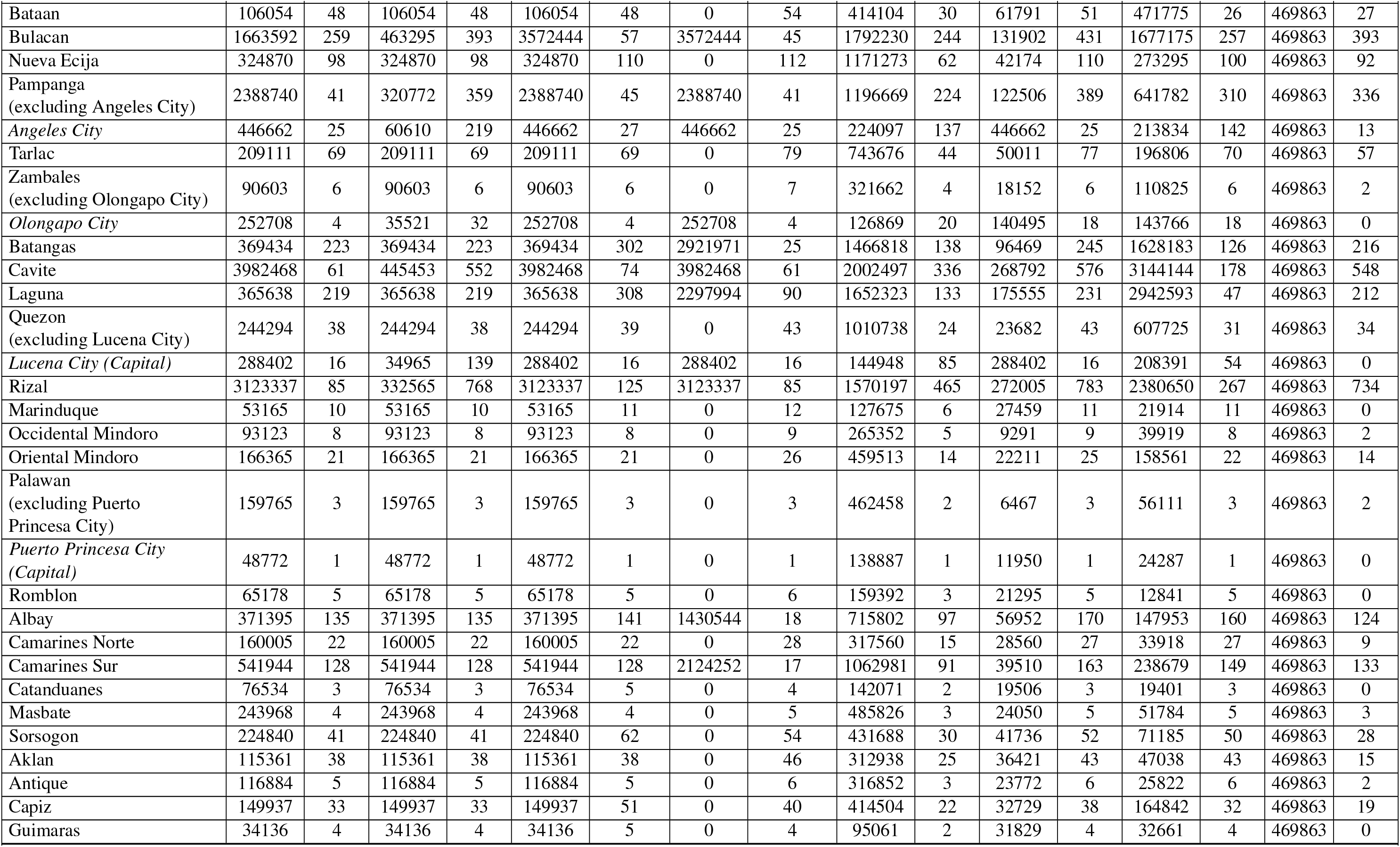
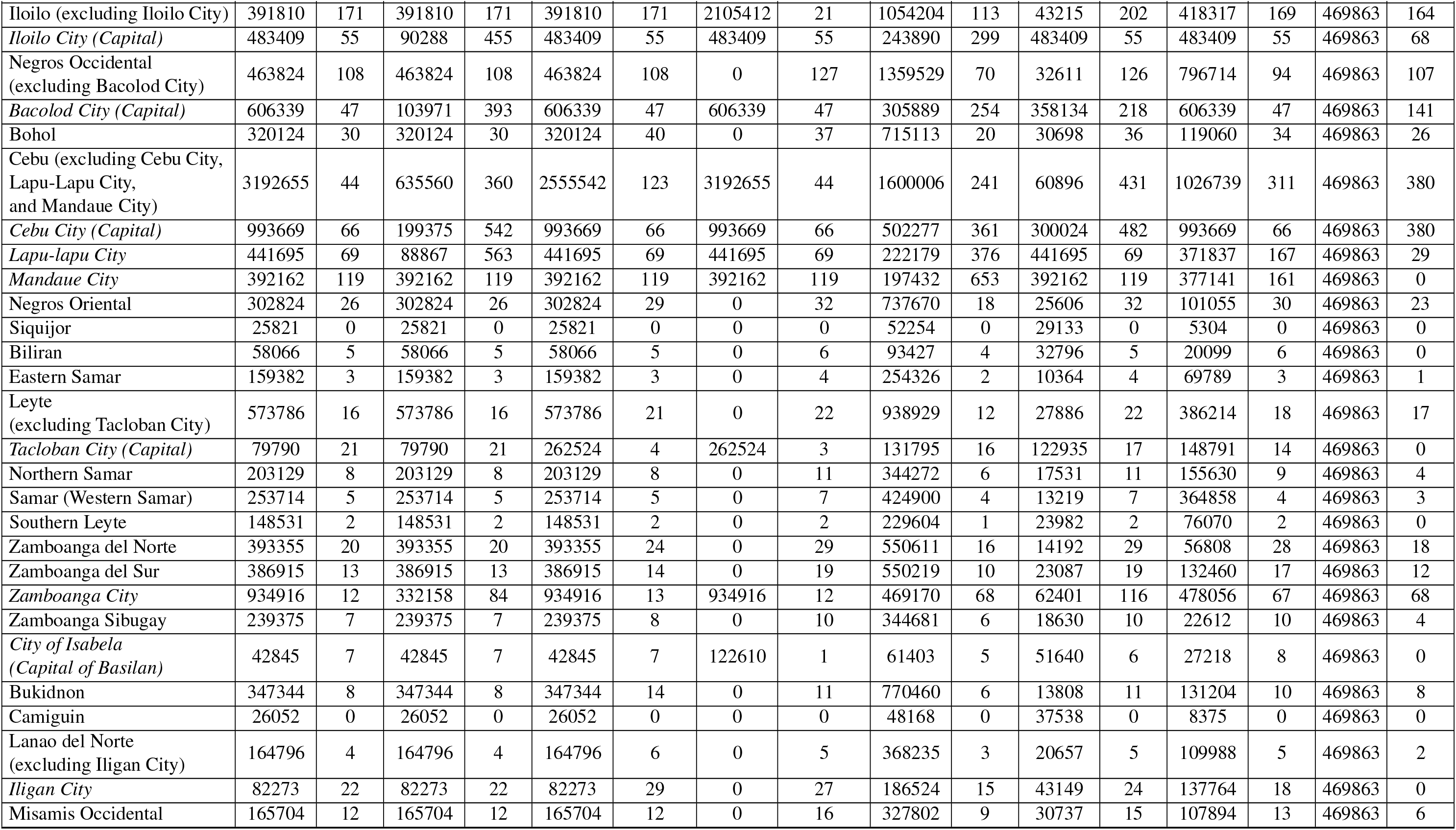
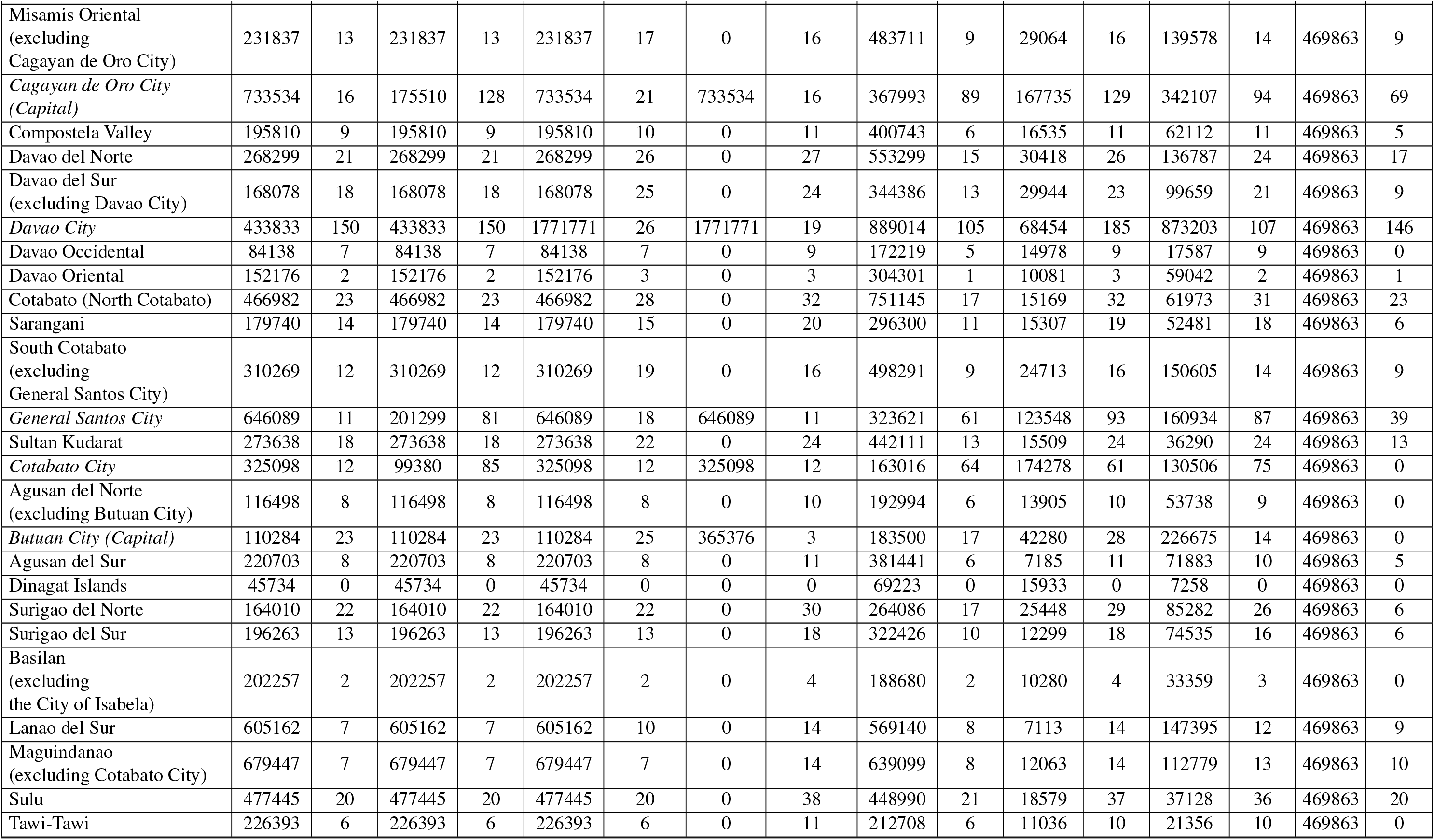
Various approaches of allocating vaccines and its resulting objective function value per city*/*province.

## Appendix D

**Table 7:** Table of Parameters

| Parameter | Description |
| --- | --- |
| $N_i$ | projected 2020 population size of locality $i$ |
| $C_i$ | number of recovered/ expired COVID-19 cases in locality $i$ as of 11/10/2020 |
| $m_i$ | number of vaccinated individuals in locality $i$ |
| eff | effectiveness rate of the vaccine |
| $R_{0i}$ | maximum reproduction number recorded in locality $i$ as of 11/11/2020 |
| $\delta_i$ | population density of locality $i$ |
| $d_i$ | case fatality rate per locality $i$ as of 11/10/2020 |
| $V$ | total available complete doses of vaccine for allocation across the country |
| $G_i$ | number of individuals belonging to the priority group in locality $i$ |
| $P$ | price per complete dose of vaccine |
| $B$ | total allotted budget for vaccination |

## References

[1] World Health Organization. Coronavirus disease (COVID-19) Situation Report 1 Philippines 9 March 2020. 2020. url: https://www.who.int/docs/default-source/wpro---documents/countries/philippines/emergencies/covid-19/who-phl-sitrep-1-covid-19-9mar2020.pdf?sfvrsn=2553985a_2 (visited on 12/17/2020).

[2] World Health Organization. Transmission of SARS-CoV-2: implications for infection prevention precautions. 2020. url: https://www.who.int/news-room/commentaries/detail/transmission-of-sars-cov-2-implications-for-infection-prevention-precautions (visited on 12/17/2020).

[3] Simran Preet Kaur and Vandana Gupta. “COVID-19 Vaccine: A comprehensive status report”. In: Virus research 288 (Aug. 2020). doi:10.1016/j.virusres.2020.198114.

[4] World Health Organization. WHO Director-General’s opening remarks at the media briefing on COVID-19 - 11 March 2020. 2020. url: https://www.who.int/director-general/speeches/detail/who-director-general-s-opening-remarks-at-the-media-briefing-on-covid-1911-march-2020 (visited on 12/17/2020).

[5] Anna Malindog-Uy. COVID-19 Impacts In The Philippines. 2020. url: https://theaseanpost.com/article/covid-19-impacts-philippines (visited on 01/15/2021).

[6] Christian Alvin Buhat and Steven Kyle Villanueva. “Determining the effectiveness of practicing non-pharmaceutical interventions in improving virus control in a pandemic using agent-based modelling”. In: Mathematics in Applied Sciences and Engineering 1 (Dec. 2020), pp. 423–438. doi:10.5206/mase/10876.

[7] The Straits Times. Philippines suffers worst job losses in 15 years due to Covid-19 and lockdown. 2020. url: https://www.straitstimes.com/asia/se-asia/philippines-suffers-worst-job-losses-in-15-years-due-to-covid-19-and-lockdown (visited on 01/15/2021).

[8] E. Murakami, S. Shimizutani, and E. Yamada. “Projection of the Effects of the COVID-19 Pandemic on the Welfare of Remittance-Dependent Households in the Philippines”. In: EconDisCliCha 288 (Sept. 2020). doi:10.1007/s41885-020-00078-9.

[9] Philippine Statistics Authority. Philippine GDP posts −8.3 percent in the fourth Quarter 2020; −9.5 percent for full-year 2020. 2021. url: https://psa.gov.ph/content/philippine-gdp-posts-83-percent-fourth-quarter-2020-95-percent-full-year-2020 (visited on 02/08/2021).

[10] Ben de Vera. COVID-19 impact: IMF sees PH GDP dropping by record 8.3 percent in 2020. 2020. url: https://business.inquirer.net/309494/covid-19-impact-imf-sees-ph-gdp-dropping-by-record-8-3-percent?fbclid=IwAR1BQZKqdp2CV3QV5nUEsqSg1ygegLmqRygjIcade5cwJGFcAa (visited on 01/15/2021).

[11] World Health Organization. International Travel and Health 2020. isbn: 978 92 4 158 047 2.

[12] Mondher Toumi and Waltere Ricciardi. “The Economic Value of Vaccination: Why Prevention is Wealth”. In: Journal of market access health policy 3 (2015). doi:10.3402/jmahp.v3.29414.

[13] Centers for Disease Control and Prevention. Vaccine Testing and the Approval Process. 2014. url: https://www.cdc.gov/vaccines/basics/test-approve.html (visited on 01/15/2021).

[14] World Health Organization. Manufacturing, safety and quality control of vaccines. 2020. url: https://www.who.int/news-room/feature-stories/detail/manufacturing-safety-and-quality-control(visited on 01/15/2021).

[15] World Health Organization. Science in 5 on Covid-19 - Episode 5. 2020. url: https://www.who.int/emergencies/diseases/novel-coronavirus-2019/media-resources/science-in-5/episode-5 (visited on 01/15/2021).

[16] CNN Philippines. PH not yet ready to handle Pfizer COVID-19 vaccine doses by January – Romualdez. 2020. url: https://cnnphilippines.com/news/2020/12/21/PH-not-yet-ready-to-handle-Pfizer-COVID-19-vaccine-doses-by-JanuaryRomualdez.html (visited on 01/15/2021).

[17] Vivienne Gulla. Philippines expects to roll out COVID-19 vaccines first half of 2021. 2020. url: https://news.abs-cbn.com/spotlight/12/10/20/philippines-expects-to-roll-out-covid-19-vaccines-first-half-of-2021?fbclid=IwAR36-JN09LR3ltspSd0nP8Wzw81PnEBsjvCz9i4k72-sh739_0 (visited on 01/15/2021).

[18] Philippine Statistics Authority. Philippine Population Density (Based on the 2015 Census of Population). 2020. url: https://psa.gov.ph/content/philippine-population-density-based-2015-census-population (visited on 04/11/2020).

[19] Department of Health. 2020. url: https://doh.gov.ph/covid19tracker x(visited on 11/25/2020).

[20] World Health Organization. COVID-19 - Immunity after recovery from COVID-19. 2020. url: https://www.who.int/emergencies/diseases/novel-coronavirus-2019/media-resources/science-in-5/episode-18---covid-19---immunity-after-recovery-from-covid-19 (visited on 02/11/2020).

[21] Kate Bubar et al. “Model-informed COVID-19 vaccine prioritization strategies by age and serostatus”. In: Science (Jan. 2021), eabe6959. doi:10.1126/science.abe6959.

[22] Matthew Hartfield and Samuel Alizon. “Introducing the Outbreak Threshold in Epidemiology”. In: PLoS pathogens 9 (June 2013), e1003277. doi:10.1371/journal.ppat.1003277.

[23] University of the Philippines. 2020. url: https://endcov.ph/statistics(visited on 11/25/2020).

[24] David Adam. A guide to R — the pandemic’s misunderstood metric. 2020. url: https://www.nature.com/articles/d41586-020-02009-w (visited on 02/08/2021).

[25] World Health Organization. COVID-19 – a global pandemic What do we know about SARS-CoV-2 and COVID-19? 2020. url: https://www.who.int/docs/default-source/coronaviruse/risk-comms-updates/update-28-covid-19-what-we-know-may-2020.pdf?sfvrsn=ed6e286c_2 (visited on 02/08/2021).

[26] Hao Hu, Karima Nigmatulina, and Philip Welkhoff. “The Scaling of Contact Rates with Population Density for the Infectious Disease Models”. In: Mathematical biosciences 244 (May 2013). doi:10.1016/j.mbs.2013.04.013.

[27] Christian Alvin Buhat et al. “Optimal Allocation of COVID-19 Test Kits Among Accredited Testing Centers in the Philippines”. In: Journal of Healthcare Informatics Research (Nov. 2020). doi:10.1007/s41666-020-00081-5.

[28] S Sanche et al. “High contagiousness and rapid spread of severe acute respiratory syndrome coronavirus 2”. In: Emerg Infect Dis 2 (2020).

[29] Department of Health. FAQS: Vaccines. 2020. url: https://doh.gov.ph/faqs/vaccines#:~:text=Frontline%5C%20health%5C%20workers%5C%2C%5C%20senior%5C%20citizens,9. (visited on 01/21/2020).

[30] Sarocha Chootipongchaivat et al. “Vaccination program in a resource-limited setting: A case study in the Philippines”. In: Vaccine 34 (Aug. 2016). doi:10.1016/j.vaccine.2016.08.014.

[31] Department of Budget and Management. GAA 2021. 2021. url: https://www.dbm.gov.ph/index.php/budget-documents/2021/general-appropriations-act-fy-2021 (visited on 02/11/2020).

[32] Katrina Domingo. Which COVID-19 vaccine is most cost-friendly for PH government? 2020. url: https://news.abs-cbn.com/news/12/10/20/which-covid-19-vaccine-is-most-cost-friendly-for-ph-government (visited on 12/10/2020).

[33] Leila B. Salaverria Julie M. Aurelio. DOH: 50 % vaccine efficacy is OK. 2020. url: https://newsinfo.inquirer.net/1376429/doh-50-vaccine-efficacy-is-ok (visited on 12/27/2020).

[34] Sarah Bartsch et al. “Vaccine Efficacy Needed for a COVID-19 Coronavirus Vaccine to Prevent or Stop an Epidemic as the Sole Intervention”. In: American Journal of Preventive Medicine 59 (July 2020). doi:10.1016/j.amepre.2020.06.011.

[35] Apoorva Mandavilli. Here’s Why Vaccinated People Still Need to Wear a Mask. 2020. url: https://www.nytimes.com/2020/12/08/health/covid-vaccine-mask.html (visited on 02/03/2021).

[36] Paul Fine, Ken Eames, and David Heymann. ““Herd Immunity”: A Rough Guide”. In: Clinical infectious diseases : an official publication of the Infectious Diseases Society of America 52 (Apr. 2011), pp. 911–6. doi:10.1093/cid/cir007.

